# Preferences for treatment for latent tuberculosis infection in primary care among people in the United States at increased risk of tuberculosis: a pilot survey

**DOI:** 10.64898/2026.05.20.26352199

**Authors:** Hélène E. Aschmann, Amy S. Tang, Meagan Lee, Katya L. Salcedo, Matthew T. Murrill, Gina Chen, Yuqian Ouyang, Kit Lui, Mehabuba Rahman, Jennifer Flood, Andrew D. Kerkhoff, Tracy K. Lin, Priya B. Shete, the Tuberculosis Epidemiologic Studies Consortium

## Abstract

**Objectives:** Tuberculosis (TB) in the United States disproportionately affects non-U.S.–born individuals. While testing this population for TB infection is recommended, little is known about individuals’ willingness to take treatment for latent TB infection (LTBI). To address this gap, we conducted a pilot preference survey among individuals from countries with high TB incidence.

**Design:** Cross-sectional survey supported by language concordant community health workers.

**Setting:** Federally qualified health center, serving a primarily Asian immigrant community, in San Francisco.

**Participants:** Adults eligible for risk-based LTBI testing based on place of birth seeking primary care.

**Outcome measures:** Perspectives on TB disease, risk of reinfection, and willingness to accept treatment if diagnosed with LTBI conditional on different factors, such as side effects, costs, and other treatment burden.

**Results:** Among 60 participants, the median age was 48 years (interquartile range 35-63 years), 52% were women, and 100% spoke Chinese. Infecting others (n=35, 58%), risk of death (n=30, 50%), and potential isolation (n=25, 42%) were the most worrisome consequences of TB disease. Reinfection risk, risk of liver damage, cost, TB progression risk, clinic visits, and blood draws were most often considered moderately or very important when deciding whether to take LTBI treatment (n=53 to 57, 88-95%). While most participants (n=56, 93%) were willing to take treatment if diagnosed with LTBI even at a 10-year TB progression risk below 1% and willing to accept a risk of liver damage (n=41, 68%), less than half would accept LTBI treatment if there were any associated cost (n=28, 47%). Finally, many participants had concerns about their reinfection risk after completing LTBI treatment (n=34, 57%).

**Conclusions:** Amongst surveyed participants, TB disease and its consequences such as hospitalization, death and infecting others were worrisome, and participants had a high level of willingness to take treatment if diagnosed with LTBI. Future assessments of how people weigh tradeoffs regarding LTBI diagnosis and treatment could inform interventions to increase LTBI treatment acceptance and completion.

## Introduction

To achieve tuberculosis (TB) elimination in the United States, broad populations must be treated for latent TB infection (LTBI).^1^ People with LTBI are infected with *M. tuberculosis* and do not have symptoms, but are at risk of developing TB disease. Non-U.S.–born communities are disproportionately affected by LTBI, with 76% of TB diagnoses in the United States occurring among these communities.^2^ LTBI treatment is effective at preventing TB disease. Despite recommendations to increase testing and treatment for LTBI among persons with risk factors,^3^ large gaps persist in the TB preventive care cascade.^4^ The recommendation for treatment and completion of treatment have been reported as steps in the cascade with greater losses.^4^ To reduce these gaps and understand whether public health goals of eliminating TB are feasible and compatible with individual decisions among people at risk of developing TB,^1^ it is crucial to understand under which conditions individuals would accept therapy for LTBI. Although some preference studies compared regimen features in diverse settings,^5–8^ little is known on how characteristics related to persons influence preferences for accepting LTBI treatment, such as presumed progression risk or whether the person would need to modify or stop a current medication.

Studies among migrants have identified multiple barriers for LTBI treatment initiation and adherence.^9–11^ The burden of LTBI treatment includes taking pills, duration of treatment, clinic visits and blood tests, as well as changes to other medications due to drug-drug interactions of rifamycins, willingness to cease alcohol consumption during the treatment, and potential costs of LTBI care, including loss of income and opportunity costs.^9,12,13^ In addition, the benefits of LTBI treatment (i.e., preventing TB) may be limited in time by a risk of reinfection, particularly if individuals travel to countries with higher TB incidence.^14^ On the other hand, individuals need to weigh the burden of LTBI treatment against the risk of developing TB and its consequences: increased risk of death,^15^ hospitalization,^16,17^ significant symptoms, stigma, isolation, and long treatment of at least 4 or 6 months.^18^ Finally, many TB survivors experience post-TB lung disease, reducing quality of life long-term and increasing mortality.^19^

To assess which of these factors and consequences exerts the most influence on LTBI treatment-related decisions, we conducted a pilot survey of non-U.S.–born persons eligible for LTBI testing. The survey (1) elicited which explanations of TB and its consequences resonate most with patients, and (2) assessed how different factors potentially affect the acceptability of LTBI treatment. These data will inform the development of a large, choice-based survey to precisely quantify trade-offs individuals are willing to accept for LTBI treatment to inform clinical practice for primary care and person-centered interventions (e.g., decision-making tools or provider education) to address gaps in the LTBI cascade of care.

## Methods

### Setting and participants

We conducted a cross-sectional pilot survey among individuals eligible for LTBI testing in October 2023. The study took place at one of the San Francisco clinic locations of North East Medical Services, a federally qualified community health center serving a large low-income Asian immigrant community.^20^ Individuals were eligible if they were born in a country with intermediate or high incidence of TB^21^ (incidence >10 per 100 000 persons), aged 18 or older, had no known prior history of TB disease (we defined individuals unsure of their prior history as eligible), and were currently receiving primary care services at North East Medical Services. Individuals were excluded if they were currently working as a health care worker or working in a hospital, nursing home, prison, correctional facility, or homeless shelter (professions with occupational LTBI testing). Individuals were also excluded if they had previously taken LTBI treatment longer than a year ago, or were unable to complete the survey in English or in Chinese (simplified or traditional), or did not provide consent. All eligibility criteria were assessed based on self-report.

### Ethics and consents

The University of California San Francisco Institutional Review Board approved this study (#23-38710). All participants provided electronic consent. This activity was reviewed by CDC, deemed not research, and conducted consistent with applicable federal law and CDC policy (see 45 C.F.R. part 46. 102 (l)).

### Survey design

We ascertained preferences to treat LTBI in the context of various clinical, social, and economic factors. An initial list of benefits, consequences of TB disease, as well as side effects from taking therapy, palatability of treatment, logistical burden of treatment, and barriers related to LTBI treatment was generated based on a review of the literature and was refined and finalized by an interdisciplinary team that included a primary care provider serving this community as well as the directors of the local Departments of Public Health TB control programs. We assessed participants’ hypothetical willingness to accept varying side effects, clinic visits, blood draws, drug–drug interactions, alcohol cessation, cost, and reinfection risk, with the importance of each factor rated using a Likert scale.

The survey also included a threshold exercise for 10-year TB risk. Participants were randomized to first see a 5% or 10% risk. The question was phrased as: “If 5 out of 100 people like you develop active TB disease in the next 10 years, would you prefer to take the treatment, or no treatment?,” or the equivalent for other risk probabilities. Based on the answer to the first question, the lowest or highest threshold was displayed next. The lowest threshold was labeled as “less than 1 out of 100” and the highest was “50 out of 100”. Further risks were shown based on the participant’s answer, proceeding to smaller risks if the answer was “yes, prefer preventive treatment” and higher risks if the answer was “no, prefer no treatment,” using the bisection method to limit the number of questions to 5 or less. All risks were displayed with an icon array (online supplemental figure S1).^22^

To rank different consequences of TB disease, participants first rated eight different consequences as not worrisome, moderately worrisome, or very worrisome. In the next step, participants were asked to identify the top three most worrisome consequences among those they rated moderately or very worrisome, and then rank these three using a drag and drop interface. The questionnaire is included in online supplemental materials 1 (English), 2 (traditional Chinese), and 3 (simplified Chinese).

### Procedures

The survey was conducted in the clinic’s lobby with the support of three community health workers who work at the community health center. The study was advertised with a poster and table. Individuals approaching the table, which was staffed by community health workers, were recruited by age, aiming for 20 participants in age groups 18-39, 40-59, and 60+ years. Participants received a $20 gift card for their participation. The survey was designed using the Qualtrics Survey Suite and was administered using an electronic tablet. Participants could complete the survey on their own or with one-on-one help from a trained, language-concordant community health worker in English, Mandarin, or Cantonese for the entire survey or as needed. All survey responses were based on self-reported data.

The survey first elicited eligibility and provided basic information on TB, LTBI, and preventive treatment for LTBI to ensure all participants had a standardized level of understanding before proceeding. Participants first ranked which consequences of TB disease would worry them the most. They then completed the threshold exercise, answered questions about potential barriers to LTBI treatment, and finally completed demographic and socioeconomic information. Willingness to accept opportunity cost was elicited in a two-step question, first asking which other activities they would prefer, and second a slider scale to elicit willingness to pay to spend time on their preferred activity instead (see questionnaire in online supplemental material 1). We randomized the order of presentation for multiple choice answers, including the consequences of TB disease, to minimize any ordering bias.

### Sample size

To describe the distribution of risk thresholds and how often other factors related to LTBI treatment could be potential barriers to treatment, we aimed for a total of 60 responses, which allowed for a frequency of 20% to be detected with a precision of +/-10% for factors related to LTBI treatment.^23^ In addition, 60 responses would allow us to detect problems (e.g., missing response options or inconsistencies) that occur in 5% of participants.^24^

### Statistical analysis

The 10-year TB progression risk threshold at which persons would prefer LBTI treatment compared with no treatment was determined as the smallest risk at which the participant responded they would prefer treatment. We used descriptive statistics to report the remaining outcomes, including the ranking of TB consequences, and the importance of different factors in the decision to take LTBI treatment or no treatment. We reported percentages calculated with a denominator of all participants, including those with missing answers, and reported the number of missing answers. Similarly, we used descriptive analyses to summarize outcomes related to willingness to accept burden or harm when taking LTBI treatment such as varying side effects, clinic visits, blood draws, drug-drug interactions, cost, and reinfection risk. In addition, for these outcomes, we classified different answers as potentially representing a barrier to LTBI care or favoring LTBI treatment (i.e., compatible with current practice) based on the investigators’ clinical experience and knowledge about the health system. All analyses were performed in R version 4.1.2.

## Results

### Recruitment and participants

Study staff were approached by 64 individuals in the clinic lobby. Four were excluded: one was U.S.– born, one was a healthcare worker, one individual did not consent (needed to go to an appointment), and one individual consented but did not complete the survey. Therefore, 60 individuals consented and completed the survey (completion rate 97% among 62 potentially eligible individuals).

Participants had a median age of 48 years (interquartile range [IQR] 35-63 years); 52% were women and 100% spoke Chinese (Table 1). Most participants were born in China, and participants immigrated a median of 15 years ago (IQR 7-23 years). 58 of 60 (97%) had health insurance. Many participants had little formal education; 52% did not complete high school or equivalent education. Two-thirds preferred not to report their household income; the majority of participants who answered reported a household income of less than $25,000. While individuals with prior LTBI testing or recent LTBI treatment were eligible, no one reported either. A few participants were unsure if they had prior testing (n=13, 22%), prior treatment (n=3, 5%), or a prior history of TB disease (n=3, 5%).

**Table 1:** Self-reported baseline characteristics of the 60 participants. For TB risk groups in the household, respondents could have multiple household members of different risk groups. IQR: interquartile range. Percentages may not add up to 100% due to rounding (education, country of birth, employment status and annual income).

|  | **Number (percent) or median [IQR]** |
| --- | --- |
| **Female** | 31 (52%) |
| **Age** | 48 [35,63] |
| **Education** |  |
| No schooling | 1 (2%) |
| Nursery school to grade 8 | 22 (37%) |
| Grade 9 or higher, without high school diploma | 8 (13%) |
| Completed high school or equivalent | 16 (27%) |
| Tertiary education | 13 (22%) |
| **Preferred language** |  |
| Chinese | 60 (100%) |
| **Country of birth** |  |
| China | 42 (70%) |
| Hong Kong | 7 (12%) |
| Vietnam | 7 (12%) |
| Taiwan | 4 (7%) |
| **Years since immigration** | 15 [7,23] |
| **Number of other people in household** | 2 [1,3] |
| **TB risk groups in household** |  |
| Senior(s) | 18 (30%) |
| Child / children | 19 (32%) |
| Immunosuppressed person | 0 (0%) |
| No risk group | 21 (35%) |
| Missing | 4 (7%) |
| **Employment status** |  |
| Employed | 28 (47%) |
| Self-employed | 4 (7%) |
| Part-time | 6 (10%) |
| Unemployed or seeking employment | 3 (5%) |
| Student or further education | 3 (5%) |
| Retired | 10 (17%) |
| Prefer not to answer | 6 (10%) |
| **Annual household income** |  |
| Less than \$25'000 | 17 (29%) |
| \$25'001 to \$50'000 | 1 (2%) |
| \$50'001 to \$75'000 | 3 (5%) |
| \$75'001 to \$100'000 | 0 (0%) |
| \$100'001 to \$150'000 | 1 (2%) |
| More than \$150'000 | 0 (0%) |
| Prefer not to answer or missing | 38 (63%) |
| **Has health insurance** | 58 (97%) |

### Importance of different consequences of TB

Participants most often rated being contagious (n=52, 87%), the potential for isolation (n=44, 73%), and risk of death (n=42, 70%) as moderately or very worrisome consequences of TB disease (Figure 1A). This pattern was consistent with a subsequent ranking exercise, where infecting others (n=35, 58%), risk of death (n=30, 50%), and potential isolation (n=25, 42%) were ranked as the top three most worrisome (Figure 1B). Stigma (i.e., others could guess they are ill with TB) and requiring treatment for TB disease (with a 6-month standard treatment regimen) were the least worrisome consequences, ranked only by 2 participants (3%) as a top three worrisome consequence. Four participants (7%) rated all consequences as not worrisome, and 18 participants (30%) rated all consequences as either moderately or very worrisome.

**Figure 1:**
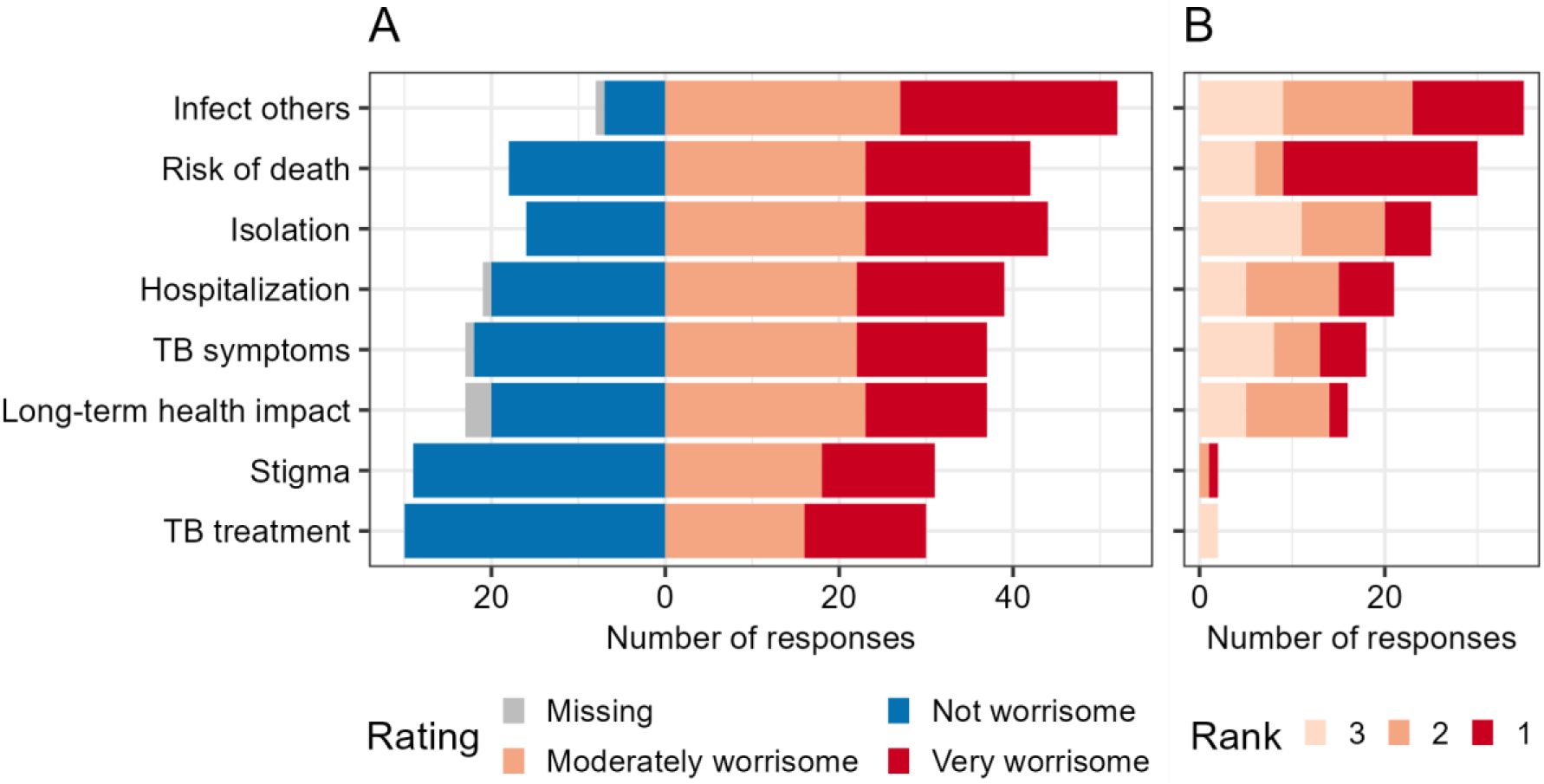
Participants rated, then ranked different consequences of tuberculosis. A) Participants rated different consequences as not worrisome, moderately worrisome, or very worrisome. B) Participants selected the top three most worrisome consequences, with rank 1 being the most worrisome. TB: tuberculosis

### Importance and acceptability of other factors

Figure 2 shows the ratings of the different factors related to the decision for or against LTBI treatment evaluated at the end of the survey. Reinfection risk (n=57, 95%), liver damage (n=57, 95%), cost or loss of income (n=56, 93%), TB progression risk (n=55, 92%), clinic visits (n=54, 90%), and blood draws (n=53, 88%) were most often rated as moderately or very important. Nausea or skin rash (n=46, 77%) and drug-drug interaction (n=44, 73%) were less often rated moderately or very important, followed by alcohol cessation (n=34, 57%). Eleven participants (18%) rated all factors for or against LTBI treatment equally, nine rated all as moderately worrisome, and two rated all as very worrisome.

**Figure 2:**
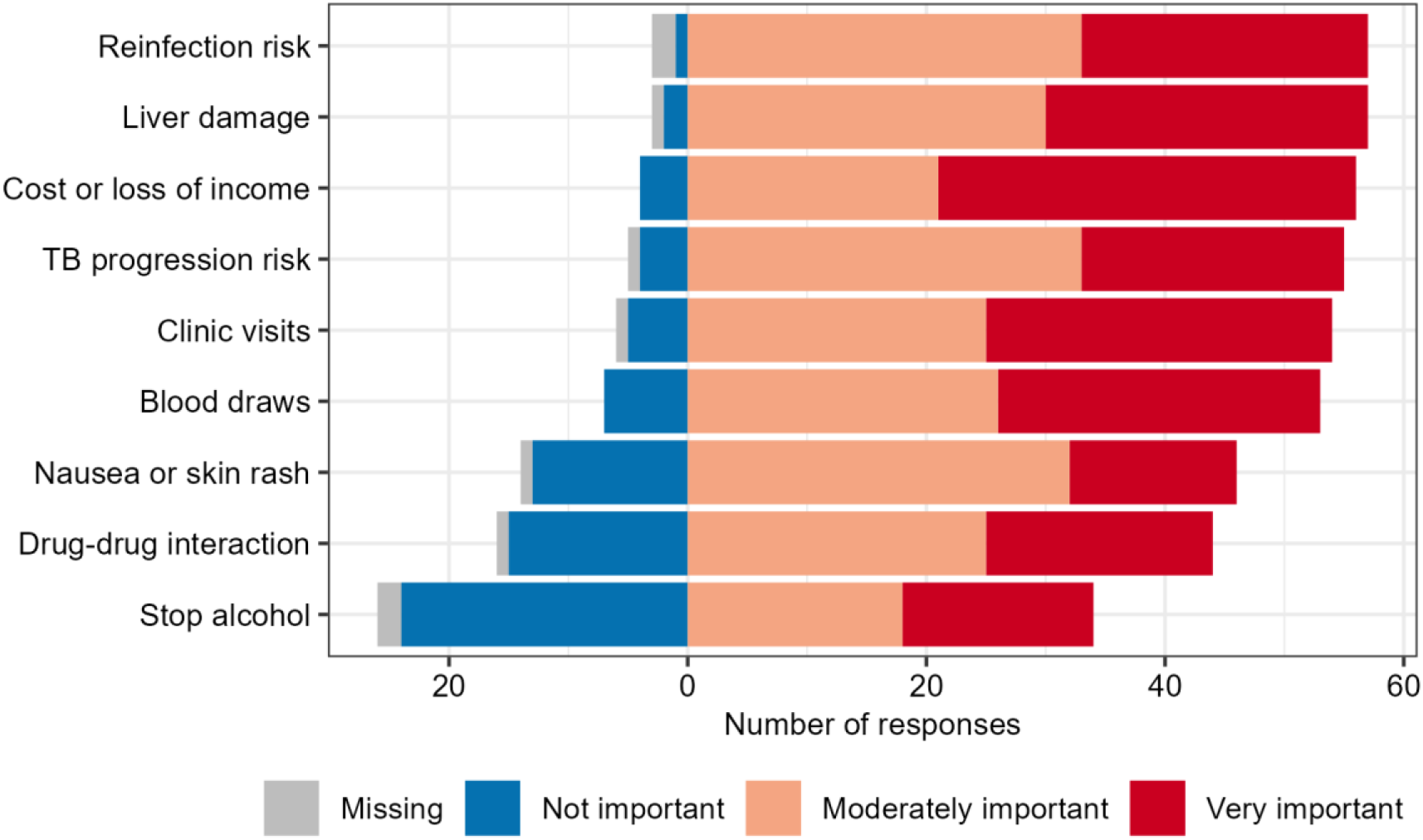
Importance of factors related to the decision for or against LTBI treatment rated on a Likert-type scale. Factors included potential barriers to treatment or burden of treatment. LTBI: latent tuberculosis infection, TB: tuberculosis

### Thresholds of TB progression risk for LTBI treatment

Almost all participants (56 of 60, 93%) indicated that they were willing to take LTBI treatment even if their risk of TB was less than 1% in 10 years (Figure 3). Three participants had risk thresholds of 3, 4, and 5%, respectively. One participant indicated not being willing to take LTBI treatment, even at a risk of 50% of developing TB in 10 years without preventive treatment.

**Figure 3:**
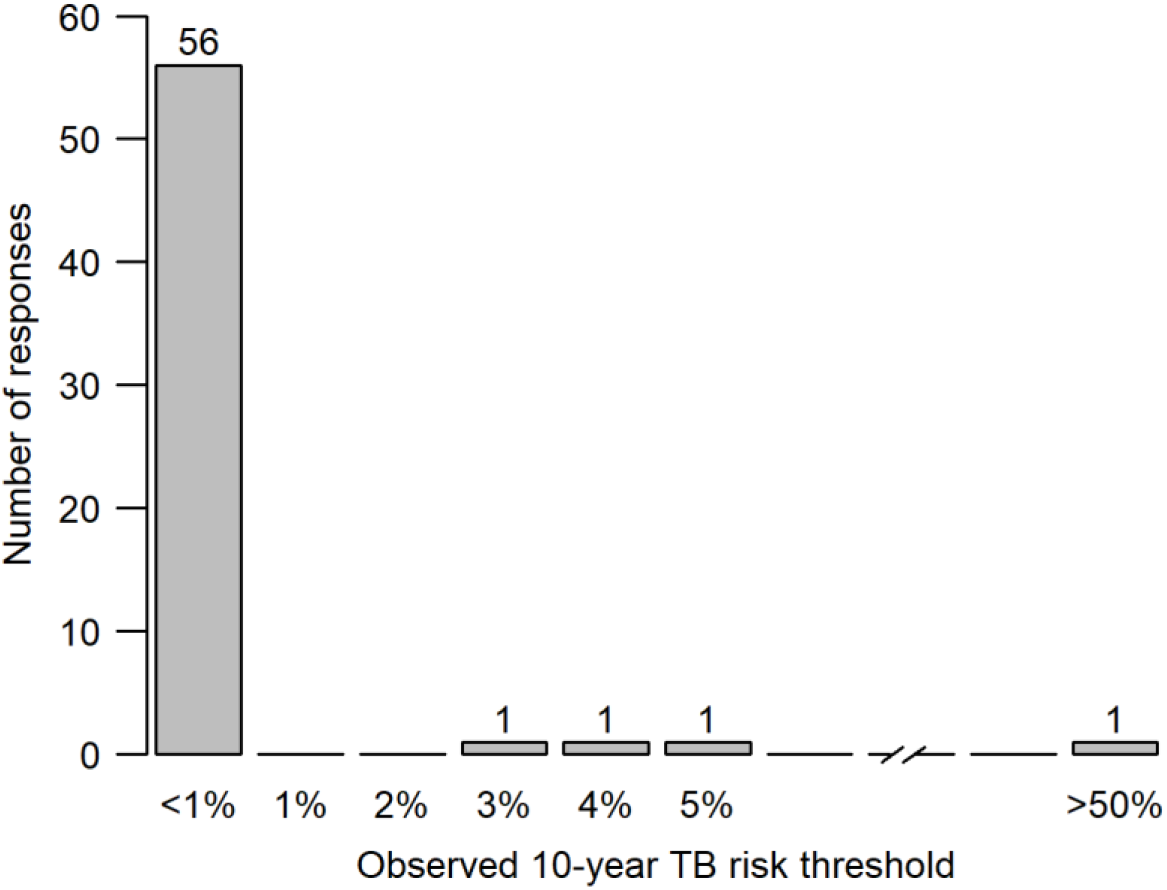
Observed tuberculosis (TB) risk thresholds after education on TB and brief education on preventive treatment

We asked participants what timeframe would be most important for when their doctor describes their risk of developing TB. Most selected one year (n=24, 40%) or lifetime risk (n=14, 23%). Some selected two years (n=9, 15%) or 5 years (n=9, 15%), and only a few selected 10 years (n=2, 3%). One did not answer, and one did not select a timeframe but specified that LTBI treatment would very likely be prescribed (i.e., the provider would recommend treatment, and they would follow the advice irrespective of risk or timeframe).

### Concern about reinfection risk

Most participants responded that they travel to countries where TB disease is common, either every year (n=8, 13%) or every 2-5 years (n=29, 48%). Only 7 (12%) did not anticipate traveling to any such country. After describing reinfection risk as about 1% among people who travel to countries where TB is common, most participants were at least slightly concerned (n=21, 35%) or reported being moderately concerned (n=12, 20%) or concerned (n=1, 2%). Less than half (n=26, 43%) were not concerned that they could “catch the TB germs again after completing preventive treatment, when traveling or for other reasons.” No one was extremely concerned.

### Willingness to accept burden, harm, and cost for LTBI treatment

#### Cost and loss of income

Figure 4 summarizes our findings ordered by how frequently factors favored no LTBI treatment (shown in red and orange) and how often participants’ values were compatible with or favored LTBI treatment (shown in blue). More than half of the participants were only willing to take TPT if it was free (n=32, 53%) and would only take LTBI treatment if they did not lose any income (n=36, 60%). Participants reported a median of $18 (IQR $0-100) as acceptable opportunity cost, described as the value of spending time on a preferred activity instead of seeking care for TB prevention.

**Figure 4:**
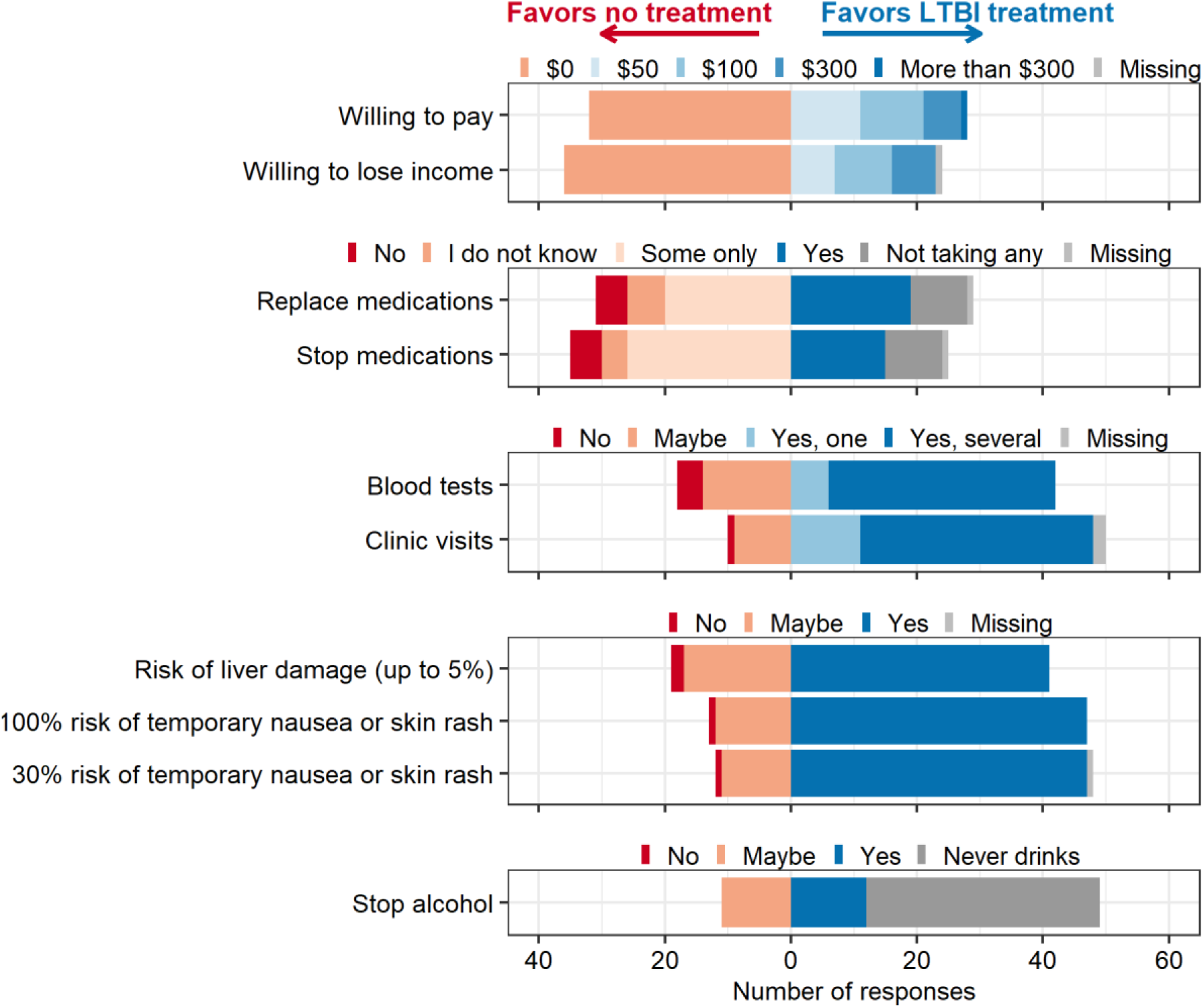
Number of participants with responses that favor LTBI treatment or favor no treatment (potential barriers). Answers that favor no LTBI treatment are shown in orange and red, answers that favor LTBI treatment, i.e., are compatible with taking treatment in the current health system, are shown in blue. Participants with missing answers or for whom the question did not apply are shown in grey. We assessed willingness to pay (out-of-pocket costs, travel costs, or cost for child / elder care), willingness to accept loss of income, willingness to accept changes to other medications for a few months, willingness to accept monthly blood tests and several clinic visits, willingness to take LTBI treatment despite risk of adverse events, and willingness to accept alcohol cessation for a few months, in order to take LTBI treatment. Monthly blood tests were presented as used to check if the liver and kidney are healthy, and LTBI treatment needs to be stopped. Clinic visits were presented as recommended to ensure the patient feels well and to help complete LTBI treatment. Temporary nausea or skin rash was described as one week, disappearing on their own without the need to see a doctor. Liver damage was described as requiring a hospital visit. LTBI: latent tuberculosis infection

#### Clinic visits and blood tests

Most participants (n=36, 60%) reported they would accept monthly blood tests, described as “to make sure my liver and kidney are healthy”, although many remained unsure (n=14, 23%, shown as “maybe” in Figure 4). Others answered that they would only accept one blood test (n=6, 10%), or that they would take LTBI treatment but not accept blood tests (n=4, 7%).

Most participants answered that they would accept and attend several clinic visits (n=37, 64%) or one clinic visit (n=11, 19%) (Figure 4). Only one participant answered they would not accept clinic visits. Among proposed visit types (in-person, by video, or phone call) with their physician or pharmacists, two visit types were strongly preferred: 37 (63%) preferred an in-person visit with their physician, and 15 (25%) preferred a phone call with their physician. Less participants preferred a video call with their physician (n=3), or visits with their pharmacist (n=1 for in-person, n=1 for video and n=2 for phone call). One participant did not rank visit types. Participants who preferred a phone call were younger (median age 39 years) than participants who preferred an in-person visit (median age 59 years).

#### Drug-drug interactions

Despite comparatively low ratings of the importance of drug-drug interactions (Figure 2), 51 of 60 participants (85%) reported taking medications: 31 took medications prescribed by their doctor, 9 took herbal or traditional medicine, 7 took over-the-counter medicine, and 5 took hormonal contraception. Among participants who took medications, the majority were only willing to stop some (n=26, 51%) or none (n=5, 10%) of their medicines for a few months if they needed LTBI treatment. Similarly, most were only willing to replace some (n=20, 39%) or none (n=5, 10%) of their medicines (Figure 4).

#### Adverse events

Liver damage was among the factors with the most participants (n=57, 95%) rating moderately or very important in the decision for or against LTBI treatment (Figure 2), whereas temporary nausea or skin rash was less often rated important (n=46, 77%). In a separate question, 20 participants (33%) reported worrying more about liver damage, 24 (40%) reported worrying about both liver damage and nausea or skin rash, and 14 (23%) worried about neither liver damage nor nausea or skin rash, as preventing TB was more important. Only 2 (3%) were more worried about nausea or skin rash than liver damage. When asked whether “given the chance of liver damage (up to 5 in 100 people)” participants would be willing to take LTBI treatment, 41 (68%) stated yes, 17 (28%) maybe, and only 2 (3%) stated no. Similar answers were given for temporary nausea and skin rash (Figure 4). Different risks of temporary nausea and skin rash did not meaningfully change participants’ willingness to take LTBI treatment.

#### Alcohol cessation

37 (62%) of the participants reported never drinking alcohol. Others reported drinking alcohol monthly or less (n=16, 27%), 2-4 times a month (n=5, 8%), and 2-3 times a week (n=2, 3%). No one reported drinking alcohol 4 or more times a week. Alcohol cessation was rated as the least important overall factor for deciding whether to accept LTBI treatment (Figure 2). Among those who reported drinking alcohol, 12 (58%) reported being willing to stop alcohol for a few months, and 11 (48%) answered maybe (Figure 4).

## Discussion

In this pilot study among non-U.S.–born individuals eligible for LTBI testing and receiving primary care at a large federally qualified health center in the United States, we found a high willingness to take LTBI treatment even at low risks of TB progression; however, participants indicated most potential harms or barriers to LTBI care that were presented to them were important. Notably, half of participants would not be willing to take LTBI treatment if there were any costs associated. This finding is especially relevant, because while there is no cost-sharing for patients with Medicaid in our setting, patients may not be aware that treatment is free.^25^ In addition, there may be co-pays for visits or diagnostic tests,^26^ and many patients face direct nonmedical costs such as travel or parking costs, or opportunity costs such as loss of income for LTBI care. ^11,13,27,28^

In addition, the risk of reinfection after LTBI treatment was a common concern. A prior study found that some providers consider LTBI treatment not acceptable if there is a risk of reinfection,^13^ which may influence shared decision-making discussions. We also found that the risk of infecting others, risk of death, the frequency of hospitalization, and long-term health impact were the most worrisome consequences of progressing to TB disease. Thus, information brochures might consider content on the signs and symptoms of TB alongside consequences of TB disease such as hospitalization, death, and potential to infect others. Although isolation was one of the most worrisome consequences, guidance shortening isolation in community settings now exists.^29^ Similarly, although we described a 6-month treatment for TB disease, a 4-month treatment is now available for drug-susceptible TB.^18^ Overall, our findings emphasize that inherent to the decision to take LTBI treatment requires patients in primary care to imagine a series of potential trade-offs that may be better considered after some amount of education and shared decision-making approach with their provider.

Among the factors we considered for LTBI treatment decision-making, alcohol cessation, mild side effects like nausea and skin rash, and drug-drug interactions were less important to participants. However, despite the comparatively low rating for the importance of drug-drug interactions, most participants reported taking other medications. Importantly, many were not willing to change or stop their medications. Therefore, drug-drug interactions and resulting medication changes may still represent a challenge to implementing TB prevention. The importance of drug-drug interactions to individuals has previously been shown for people living with HIV.^6,30^ Unfortunately, the shorter, preferred drug regimen contain rifamycins, which often interact with other drugs. Therefore, a regimen of 6 months of isoniazid is recommended for people unable to take a shorter preferred regimen due to drug intolerability or drug-drug interactions with rifamycins.^31^ Consistent with the overall importance rating, nausea and skin rash were considered less important than the risk of liver damage (a more severe side effect). This preference pattern is consistent with results from a study in Canada, where participants were more concerned about liver damage than skin rashes or fatigue,^5^ and a study in South Africa and one in Uganda, where side effects were least prioritized compared to other factors such as preventing disease.^6,32^ Liver damage may have been highly rated among individuals included in our sample, partly due to the importance of the liver in traditional Chinese medicine.^33^

Over 90% of participants were willing to take LTBI treatment even at a 10-year progression risk below 1%. The implication is that these participants would want treatment if they knew they were infected, regardless of their level of risk. These findings counter some providers’ perception that individuals do not want or prioritize LTBI care, as illustrated in one qualitative study.^13^ Prior preference studies in different settings evaluated treatment choices at higher progression risks found variable acceptability: in Canada with an option for 9 months of isoniazid with a presented 10% risk over 10 years, 75% accepted LTBI treatment^5,34^, whereas in Uganda with regimens of 1 or 3 months of isoniazid and rifapentine and a presented risk of 5% over 3 years,^6^ over 90% indicated they would accept treatment. In real-world clinical settings in the United States, however, treatment initiation rates are much lower.^20^ In addition, studies from high TB incidence settings showed that barriers to treatment uptake are reduced after TB education,^35^ and that the awareness of being at risk of TB increased treatment acceptance.^36^ Thus, our observed risk thresholds might have been low partly because participants were well-informed about TB and its consequences before completing the threshold exercise.

This study contributed to our understanding of preference for LTBI treatment amongst a non-U.S.–born Chinese-speaking population by including a detailed survey with high completion through self-administration or administration by a language-concordant community health worker. These steps allowed the broad inclusion of people of different ages and with less education. As we broadly assessed different factors for their relevance in the decisions around LTBI treatment, our findings can inform the design of a larger choice-based study, both for the selection of attributes and their levels.

The main limitations of our cross-sectional survey include the sample size (as this was only a pilot study), representativeness, and hypothetical choices. Participants were not representative of all non-U.S.–born populations, but were invited among an Chinese-speaking patient population in San Francisco with high prevalence of TB infection.^20^ Our survey measured hypothetical treatment preferences, and a larger choice-based study could better predict actual treatment choices.^34,38^ Furthermore, when completing the threshold exercise, participants had only minimal prior information from the survey about LTBI treatment, which was described as 3-4 months of daily treatment with 1-2 antibiotics. We presented this task early in the survey before discussing all potential barriers in detail because we assumed it was the most cognitively demanding. Therefore, it is possible that risk thresholds would be higher if participants knew more about the trade-offs, and future studies should evaluate the trade-offs between TB prevention and treatment burden using designs such as discrete choice experiments or conjoint analyses. In addition, the results of the rating exercise of all factors related to LTBI treatment should not be considered alone, as responses to other questions revealed that the importance could be underestimated (e.g., changes to medications) or overestimated (e.g., alcohol cessation among participants reporting no alcohol use). Furthermore, numeracy may be low in this population, as many participants had little education. We used icon arrays to facilitate risk communication and address potential low numeracy.^22,37^ Moreover, the survey administration by community health workers may have affected responses and led to social desirability bias.

In conclusion, through this survey we found that among adults of all ages eligible for LTBI testing based on place of birth who receive primary care at a community health center, the willingness to take LTBI treatment was high even in the context of low risk of progression, as long as testing and treatment is free. However, the benefit of treatment for avoiding progression to the harms of TB disease is likely often a trade-off with potential burdens or harms, including adverse events, clinic visits, blood draws, and changes to other medications due to drug-drug interactions. Our findings can inform future studies that may measure importance of these trade-offs with choice-based designs, ideally also considering whether the risk of reinfection matters to patients. Finally, our findings may inform person-centered decision-making tools for TB prevention in primary care settings.

## Data availability statement

Deidentified individual participant data that support the findings of this study are available from the University of California San Francisco upon reasonable request after approval of a proposal.

## Patient consent for publication

Not applicable

## Contributors

HEA, PBS, TKL conceived and designed the study. AST, ML, KLS, MTM, GC, YO, KL, MR, JF, ADK contributed to the design and interpretation of the results. GC, YO, KL translated the survey and recruited and surveyed participants. HEA and PBS were involved in funding acquisition. HEA analyzed the data and drafted the manuscript. PBS and TKL are responsible for the overall content as the guarantor. The authors read and approved the manuscript.

## Acknowledgements

We are grateful to the participants in this study who shared their views on tuberculosis and healthcare decision-making with us. We are grateful to the TB Research and Mentorship Program at UCSF for providing structured mentorship to help cultivate this work.

