## Supplementary material for "Preferences for treatment for latent tuberculosis infection in primary care among people in the United States at increased risk of tuberculosis: a pilot survey": Figure S1

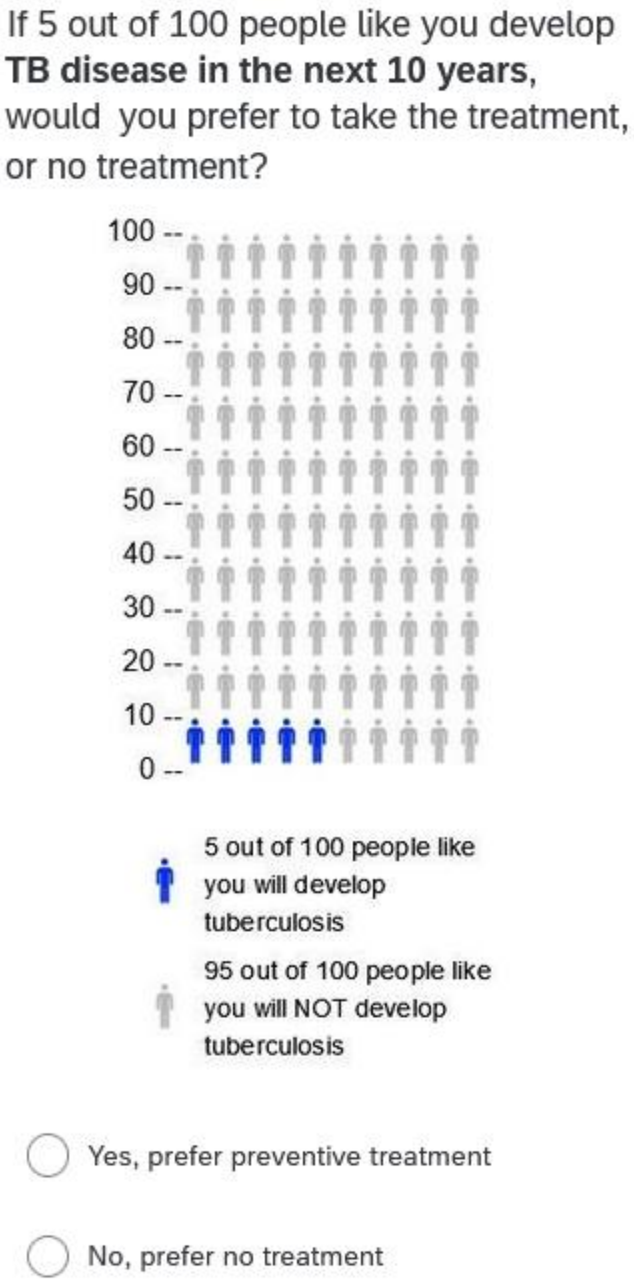


*Online Supplemental Figure S1: Screenshot of an example task from the threshold exercise with a 5% risk of progression to active TB disease.*
