## Supplementary material for "Preferences for treatment for latent tuberculosis infection in primary care among people in the United States at increased risk of tuberculosis: a pilot survey": Online Supplement 1

Online supplemental material 1

### Questionnaire in English

Start of Block: Block_initialCheck

We would like to invite you to a survey on your preferences and values regarding care for latent tuberculosis infection (latent TB). People who have **latent TB** are healthy and do not have any symptoms, but they have the tuberculosis germ. This germ is sleeping but can wake up and cause tuberculosis, which is a serious illness. First, we will ask you some questions about yourself and where you receive care to find out if you can participate in this survey.
Among the following languages, what is your preferred language to complete this survey in?

- ○ English
- ○ Traditional Chinese
- ○ Simplified Chinese
- ○ I am not comfortable completing a survey in any of the above languages

---

What is your age?

- ○ Years:________________________________________________

What is your sex?

- ○ Male
- ○ Female

[This question was modified from the original by requirement of the CDC, funders of the study, to comply with an Executive Order enacted on January 20, 2025.]

What is the highest level of education you completed?

- ○ No schooling
- ○ Nursery school to grade 8: elementary / primary / middle school
- ○ Grade 9 or higher but no high school diploma
- ○ Completed high school or equivalent, or trade/technical/vocational training
- ○ Tertiary education: college, university or higher

Do you currently receive primary care at NEMS?

- ○ Yes
- ○ No, elsewhere

*Go to Block_ineligible if age below 18, not at NEMS, or not comfortable in any of the offered language.*
*Otherwise continue to Block_CountryOrigin.*

End of Block: Block_initialCheck

Start of Block: Block_ineligible

MessageIneligible Thank you for your interest in this study and for taking time to fill out the pre-screening questions. Based on your answers, you are not eligible to be in this study. **Thank you** again for your help and participation.

*End survey.*

End of Block: Block_ineligible

Start of Block: Block_CountryOrigin

Persons who were born in a country where **TB disease** is more common are recommended to get tested for **latent TB**. Please answer a few questions about where you are from and any experience you have with **TB disease** or **latent TB** to check if you are eligible for this survey.

Were you born in the United States?

- ○ Yes
- ○ No

*Skip To: End of Block if Yes*

In which country were you born?

- ○ China
- ○ Vietnam
- ○ Hong Kong
- ○ Philippines
- ○ Taiwan
- ○ Mexico
- ○ India
- ○ Thailand
- ○ Other

*Display this question if Other*

In which other country were you born?

▼ Afghanistan ... Zimbabwe (1357)

When did you move to the United States?

- ○ Please enter year in format: YYYY________________________________________________

*Go to Block_ineligible if born in any of these countries: San Marino, Saint Kitts and Nevis, Monaco, Barbados, United Arab Emirates, Saint Lucia, United States of America, Slovakia, Israel, Norway, Iceland, Andorra, Grenada, Jamaica, Finland, Hungary, Sweden, Denmark, Czech Republic, Croatia, Greece, Jordan, Slovenia, Netherlands, Cyprus, Switzerland, Ireland, Italy, Antigua and Barbuda, Germany, Austria, Canada, Oman, Luxembourg, United Kingdom of Great Britain and Northern Ireland, Australia, Samoa, New Zealand, Cuba, Tonga, France, Belgium, Spain, Saudi Arabia, Saint Vincent and the Grenadines, Estonia, Lebanon.*

*Otherwise continue to Block_workexclusions.*

**End of Block: Block_CountryOrigin**

**Start of Block: Block_workexclusions**

Do you work as a health care worker?

- ○ Yes
- ○ No
- ○ I don't know

*Skip To: End of Block if Yes*

Do you work in a hospital, nursing home, prison, correctional facility, or homeless shelter?

- ○ Yes
- ○ No
- ○ I don't know

*Go to Block_ineligible if yes for any of these professions.*
*Otherwise continue to Block_TBhistory.*

**End of Block: Block_workexclusions**

Start of Block: Block_TBhistory

Have you ever been told by a doctor or public health department that you were sick with active tuberculosis (TB) disease?
**TB disease** is often a disease in the lung. It is a serious, and life-threatening illness. Common symptoms include coughing, fever, night-sweats, and rapidly losing weight (without trying to). If you had **TB disease**, you likely took 6 months or more of antibiotic treatment with 4 different drugs.

- ○ Yes
- ○ No
- ○ I don't know

*Skip To: End of Block if Yes, then go to Block_ineligible. Otherwise continue.*

Persons who are infected with latent tuberculosis (**latent TB**) are healthy and do not have any symptoms, but they have the TB germ. This germ is in a sleeping state.

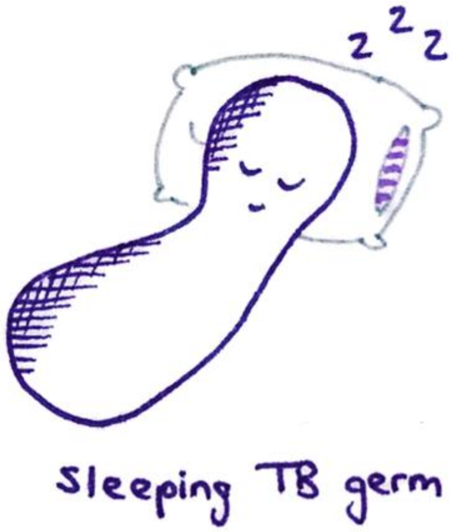


The germ can reactivate and cause **TB disease**, a serious, and life-threatening illness. People who have **latent TB** are usually recommended to take preventive treatment to make sure the germ does not reactivate.

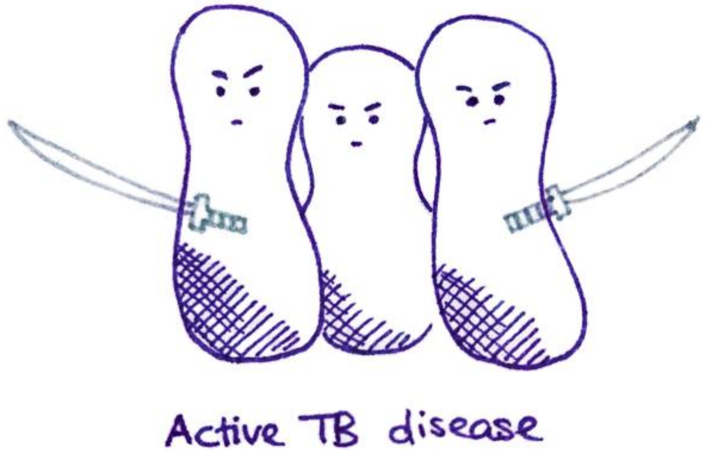

Have you ever had a test for **latent TB** infection? (a skin test or a blood test)

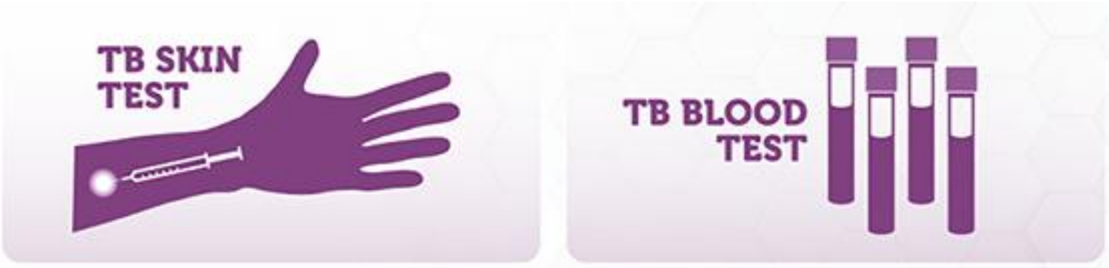


- Yes
- No
- I don't know

*Go to Block_iftested if yes.*
*Otherwise skip block and continue to BlockPriorTPT.*

End of Block: Block_TBhistory

Start of Block: Block_iftested

What type of test did you have? Check all that apply.

- ☐ Tuberculin skin test
- ☐ Blood test
- ☐ I don't know

Was any of these tests positive (meaning that you have the TB germ)?

- Yes
- No
- I did not yet receive the test result
- I don't know

*Display this question if Yes*

When did you **first** have a **positive** latent TB test?

- Longer than a year ago
- Less than a year ago

End of Block: Block_iftested

Start of Block: BlockPriorTPT

Have you ever taken preventive treatment for **latent TB** (healthy but infected with TB germ)? This is a treatment with 1-2 antibiotic medicines you take for several months to reduce your risk of TB disease.

- Yes
- No
- I don't know

*Display this question if Yes*

When did you take preventive treatment?

- I am currently taking preventive treatment
- I started preventive treatment less than a year ago and completed it or stopped.
- Longer than a year ago.

*Go to Block_ineligible if longer than a year ago.*
*Otherwise continue to BlockDigitalConsent.*

End of Block: BlockPriorTPT

Start of Block: BlockDigitalConsent

University of California
San Francisco

**Consent Form**

This is a research study, and you do not have to take part. If you have any questions, you may contact the study team members listed below. NEMS is participating in this research study with the University of California, San Francisco (UCSF). The study is led by Dr. Priya Shete at UCSF and Dr. Amy Tang at NEMS.

You are being asked to take part in this study because people who were born outside the US (from most other countries) are recommended to be screened for latent tuberculosis. People who have latent tuberculosis are healthy and have no symptoms, but they are at risk to develop active tuberculosis, a serious illness.

In this study, the researchers are doing a survey to learn more about your perspective on testing or preventive treatment for latent tuberculosis infection. The CDC (Centers for Disease Control and Prevention) is paying for this research. About 1400 people will participate in different surveys in this study.

**What will happen if I take part in this study?**

If you agree to be in this study, you will complete a survey and provide feedback on it. The survey asks about your perspective on testing or preventive treatment for latent tuberculosis infection through a series of hypothetical questions. This survey will help us learn more about when people should be tested and treated for latent tuberculosis.

Throughout the survey, we will ask you to think aloud and report if anything is difficult to understand or can be improved. We will ask you additional questions on your thoughts about latent tuberculosis and what matters most to you. The survey and interview will take about 30 minutes to 1 hour to complete. You can complete the survey on a tablet or ask the interviewer to help you enter your answers. One or two interviewers will be present and will take written notes about your feedback and reactions. Our goal is to improve the survey. You will not be audio or video recorded. The interview will take place in-person at NEMS.

**How will my information be used?**

Researchers at UCSF will use your information to inform this study. Once the study is completed, we may use de-identified information or share it with other researchers for future research studies. We will not share your name or any other personal information. We will not ask you for additional permission to share this de-identified information.

**Are there any risks to me or my privacy?**

Some of the survey questions may make you feel uncomfortable or raise unpleasant memories. You are free to skip most questions or stop the interview at any time. We will do our best to protect the information we collect from you. We will not collect any information that could identify you, such as your name or address. Authorized representatives from the University of California may review your research data for the purpose of monitoring or managing the conduct of this study.

**Are there benefits?**

There is no direct benefit to you. Although this does not help you directly, this survey will help us understand people's testing and treatment preferences and might benefit other people in the future.

**Can I say "No"?**

Yes, you do not have to participate in this interview. Your decision to complete this interview or not, will not impact your medical care at NEMS in any way.

**Are there any payments?**

When you complete this interview, you will receive a Target gift card of $20 for your participation from UCSF.

**Who can answer my questions about the study?**

Please contact Dr. Amy Tang or Dr. Priya Shete. If you have questions or concerns about your rights as a research participant, you can call the UCSF Institutional Review Board at 415-476-1814.

We need to be sure you understood this consent form:

Is your participation in this survey optional?

- ○ Yes
- ○ No

Will your participation in this interview have an impact on the care you receive at NEMS?

- ○ Yes
- ○ No

If you want to participate in this study, check "Yes" and click the "Next" button to start the survey.

Do you consent to participate in this study?

- ○ Yes
- ○ No

*Go to Block_ineligible if no consent.*
*Otherwise continue to StartSurveyC*

**End of Block: BlockDigitalConsent**

---

**Start of Block: StartSurveyC**

Who is interviewing you today?

- ○ <Community health worker 1>
- ○ <Community health worker 2>
- ○ <Community health worker 3>
- ○ < Researcher 1>
- ○ No one, I am completing this survey on my own.
- ○ Other, please specify:________________________________________________

This survey will discuss different factors that people may consider when deciding about preventive treatment for **latent TB** (healthy but infected with TB germ). We would like to understand which factors matter to you.

**End of Block: StartSurveyC**

---

**Start of Block: BlockActiveTB**

**TB disease** is a serious disease. How much would you worry about different consequences of **TB disease**? Each of these consequences represents a normal experience for someone with **TB disease** in the United States.

UCSF University of California San Francisco

| | Not worrisome | Moderately worrisome | Very worrisome |
|---|---|---|---|
| Hospitalized: You are in the hospital for 2 weeks. In the hospital, you are isolated in a room, with little contact to anyone. | ○ | ○ | ○ |
| Ill health: You cough a lot, lose weight, and have fever and night sweats. After 1 month, you find out that you have TB. The symptoms improve after 2 weeks on treatment and are gone by 2 months. | ○ | ○ | ○ |
| Quarantine: When you leave the hospital, you are still infectious and are forced to stay at home and isolate. You cannot go back to work for 2 months. | ○ | ○ | ○ |
| Treatment (6 months): You take daily pills for 6 months; 10 pills for the first 2 months, and 4 pills for the last 4 months. You often have nausea from the medication. You have regular clinic visits and a nurse checks every day that you took your pills. | ○ | ○ | ○ |
| Continued impact on health: Many people who had active TB disease recover, but never feel quite as healthy as before they had TB. | ○ | ○ | ○ |
| Infectious: Before you get diagnosed and treated, you could infect others in your household or family. | ○ | ○ | ○ |
| Stigma: Others could find out that you had TB. Because you are missing at work, isolating, and contact tracers test your colleagues and friends for TB infection, they may guess you have TB. | ○ | ○ | ○ |
| Risk of death: Some people are diagnosed too late. In California, 12% of people with TB die. | ○ | ○ | ○ |

Intro_TB_top3 What would worry you most? Please select your top 3 among the ones you found worrisome.

*Display each choice if selected as “Very worrisome” or “Moderately worrisome”*

- ▢ Hospitalized: You are in the hospital for 2 weeks. In the hospital, you are isolated in a room, with little contact to anyone.
- ▢ Ill health: You cough a lot, lose weight, and have fever and night sweats. After 1 month, you find out that you have TB. The symptoms improve after 2 weeks on treatment and are gone by 2 months.
- ▢ Quarantine: When you leave the hospital, you are still infectious and are forced to stay at home and isolate. You cannot go back to work for 2 months.
- ▢ Treatment (6 months): You take daily pills for 6 months; 10 pills for the first 2 months, and 4 pills for the last 4 months. You often have nausea from the medication. You have regular clinic visits and a nurse checks every day that you took your pills.
- ▢ Continued impact on health: Many people who had active TB disease recover, but never feel quite as healthy as before they had TB.
- ▢ Infectious: Before you get diagnosed and treated, you could infect others in your household or family.
- ▢ Stigma: Others could find out that you had TB. Because you are missing at work, isolating, and contact tracers test your colleagues and friends for TB infection, they may guess you have TB.
- ▢ Risk of death: Some people are diagnosed too late. In California, 10% of people with TB die, mostly because they are diagnosed too late.

Intro_TB_ranking What would worry you most? Please rank using drag and drop (1: most worrisome, 3: least worrisome).

*Display each choice if selected among top three*

______ Hospitalized: You are in the hospital for 2 weeks. In the hospital, you are isolated in a room, with little contact to anyone.
______ Ill health: You cough a lot, lose weight, and have fever and night sweats. After 1 month, you find out that you have TB. The symptoms improve after 2 weeks on treatment and are gone by 2 months.
______ Quarantine: When you leave the hospital, you are still infectious and are forced to stay at home and isolate. You cannot go back to work for 2 months.
______ Treatment (6 months): You take daily pills for 6 months; 10 pills for the first 2 months, and 4 pills for the last 4 months. You often have nausea from the medication. You have regular clinic visits and a nurse checks every day that you took your pills.
______ Continued impact on health: Many people who had active TB disease recover, but never feel quite as healthy as before they had TB.
______ Infectious: Before you get diagnosed and treated, you could infect others in your household or family.
______ Stigma: Others could find out that you had TB. Because you are missing at work, isolating, and contact tracers test your colleagues and friends for TB infection, they may guess you have TB.
______ Risk of death: Some people are diagnosed too late. In California, 10% of people with TB die, mostly because they are diagnosed too late.

Page Break

People who have **latent TB** (healthy but infected with TB germ) may or may not develop **TB disease**. How likely **TB disease** develops in a person depends on their age, smoking status, and general health, and other factors. The Centers for Disease Control and Prevention (CDC) recommends TB preventive treatment.
However, some people prefer to make a trade-off between preventing **TB disease** and 3-4 months of daily treatment. If people are likely to develop **TB disease**, they may benefit more from TB preventive treatment. If people are unlikely to develop **TB disease**, taking TB preventive treatment has only a small benefit of making it even less likely. We are interested to learn at what risk of developing **TB disease** you would take preventive treatment.

For the next questions, please imagine that you have **latent TB**. You have a chance of developing **TB disease** in the **next 10 years**.

*Randomize to BlockThreshold5 or BlockThreshold10, i.e., starting with 5% or 10% risk of TB disease. Only show one of the two blocks.*

**End of Block: BlockActiveTB**

**Start of Block: BlockThreshold5**

If 5 out of 100 people like you develop **TB disease in the next 10 years**, would you prefer to take the treatment, or no treatment?

- ○ Yes, prefer preventive treatment
- ○ No, prefer no treatment

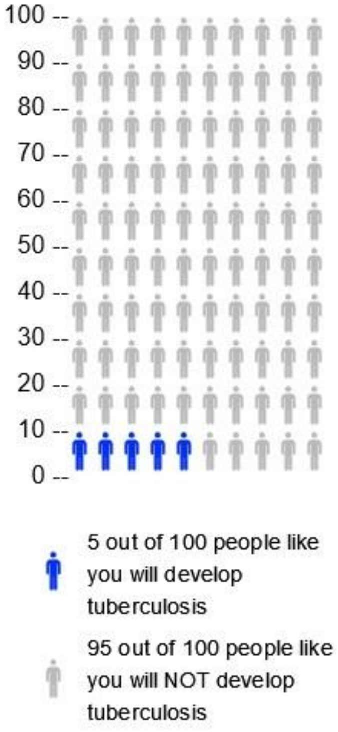


*If yes, skip to "less than 1 out of 100". If no, continue to 50%. Follow the logic shown below. Arrows to the left mean "yes", arrows to the right mean "no". A straight arrow down to "stop" means go to the End of Block, regardless of the answer.*

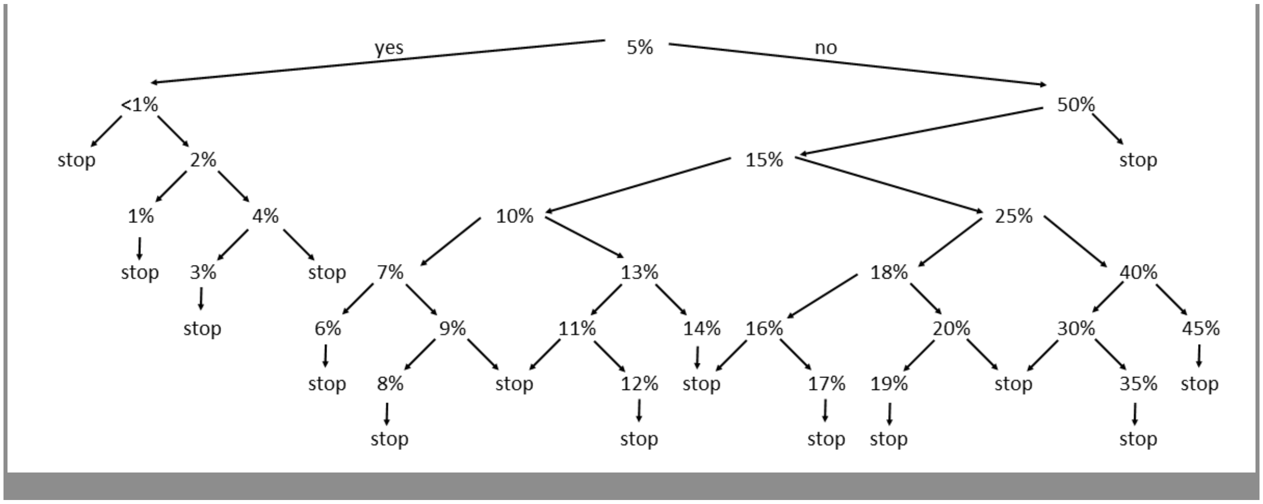


*At the end of this block, skip to Block_TBrisk_sliders.*

**End of Block: BlockThreshold5**

**Start of Block: BlockThreshold10**

If 10 out of 100 people like you develop **TB disease in the next 10 years**, would you prefer to take the treatment, or no treatment?

○ Yes, prefer preventive treatment
○ No, prefer no treatment

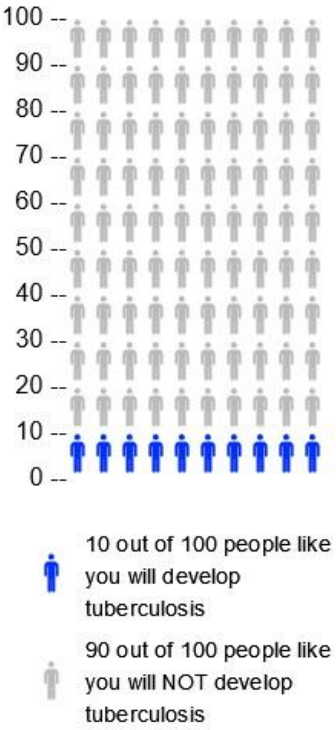


*If yes, skip to “less than 1 out of 100”. If no, continue to 50 out of 100. Follow the logic shown below. Arrows to the left mean “yes”, arrows to the right mean “no”. A straight arrow down to “stop” means go to the End of Block, regardless of the answer.*

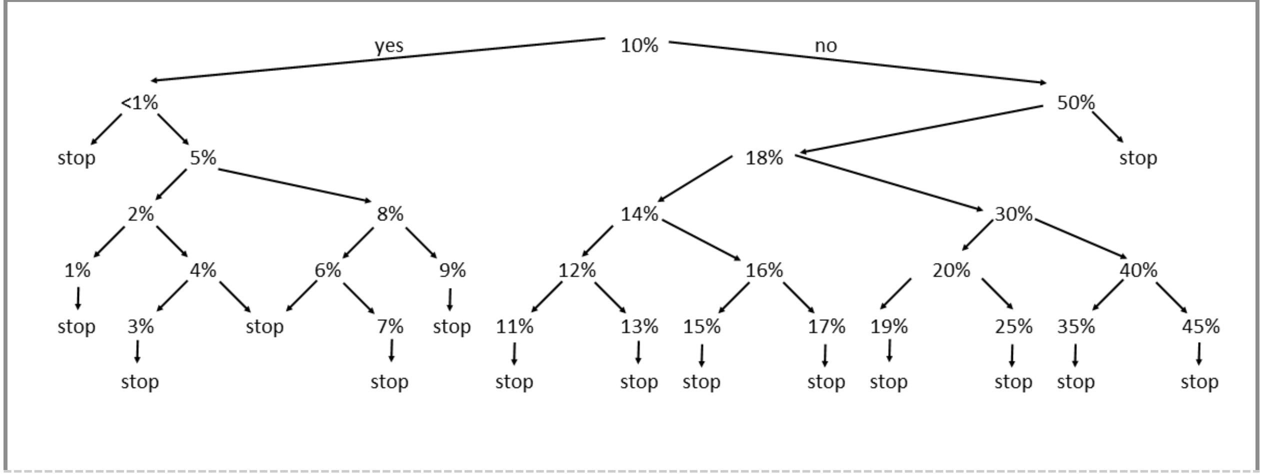


[We acknowledge that there was a translation error in the legend of the icon array for the risk below 1%. However, the question was translated and administered correctly, and we are confident the question was interpreted correctly by all participants.]

*At the end of this block, skip to Block_TBrisk_sliders.*

End of Block: BlockThreshold10

Start of Block: Block_TBrisk_sliders

We will now discuss how time impacts your thoughts about wanting TB preventive treatment.
[randomized order of questions: 10 years, 2 years, life time risk]

Consider your chance to develop **TB disease in the next 10 years**.
For you to take preventive treatment, how high would your chance of getting TB disease need to be? (For example, think which chance would be too small?)  Click on the scale to answer.

| | Low chance (%) | High chance (%) |
|---|---|---|
| | 0 10 20 30 40 50 | 60 70 80 90 100 |
| Chance (%) to develop TB in the **next 10 years** () | | |

If you were to develop **TB disease**, it would likely happen in the **next 2 years**.
For you to take preventive treatment, how high would your chance of getting TB disease need to be? (For example, think which chance would be too small?)

| | Low chance (%) | High chance (%) |
|---|---|---|
| | 0 10 20 30 40 50 | 60 70 80 90 100 |
| Chance (%) to develop TB in the **next 2 years** () | | |

TBslider_lifetime Consider your chance to develop **TB disease over your entire life-time**. For example, if you expect to live up to 90 years, consider your chance to develop **TB disease** until age 90.  For you to take preventive treatment, how high would your chance of getting **TB disease** need to be? (For example, think which chance would be too small?)

| | Low chance (%) | High chance (%) |
|---|---|---|
| | 0 10 20 30 40 50 | 60 70 80 90 100 |
| Chance (%) to develop TB in **during your lifetime** () | | |

End of Block: Block_TBrisk_sliders

Start of Block: Block_TBrisk_sliders2

When your doctor explains your chance of developing **TB disease**, what time frame is most important to you in thinking about your chance of developing TB?

- ○ 1 year
- ○ 2 years
- ○ 5 years
- ○ 10 years
- ○ 15 years
- ○ 20 years
- ○ 50 years
- ○ Lifetime risk
- ○ Other, please specify________________________________________________

End of Block: Block_TBrisk_sliders2

Start of Block: Side effects

The TB preventive treatment sometimes may sometimes cause side effects such as nausea or skin rash for about one week. These side effects will disappear on their own.
People do not need to see a doctor.

Would you still take TB preventive treatment, even though you may experience temporary nausea or skin rash?

- ○ Yes
- ○ Maybe
- ○ No

Each person's chance of experiencing nausea or skin rash for about one week is different. Imagine 30 in 100 persons like you experience nausea or skin rash. Would you still be willing to take the TB preventive treatment?

- ○ Yes
- ○ Maybe
- ○ No

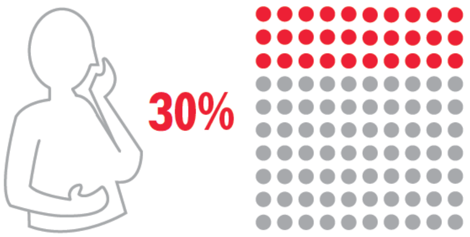

Imagine everyone would get nausea or skin rash for a week for about one week. Would you still be willing to take the TB preventive treatment?

- ○ Yes
- ○ Maybe
- ○ No

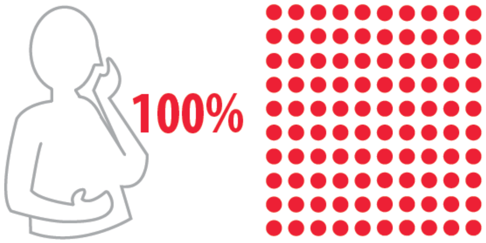


Page Break

TB preventive treatment can also cause severe side effects such as liver damage. Between 0 to 5 in 100 people will develop liver damage. When a person gets liver damage, they need to go to the hospital.

Given the chance of liver damage (up to 5 in 100 people), would you be willing to take TB preventive treatment?

- ○ Yes
- ○ Maybe
- ○ No

Do you worry more about:

- less common liver damage (up to 5 in 100 people) OR
- more common nausea or a skin rash for a week (30 in 100 people)?

- ○ I worry more about liver damage.
- ○ I worry more about nausea or skin rash.
- ○ I have the same worry about both.
- ○ I do not worry about liver damage, nausea or skin rash. Preventing TB is more important to me.
- ○ Other, please specify:________________________________________________

Do you have any comments about side effects of TB preventive treatment? (Skip if no comments.)

______________________________________________________________

End of Block: Side effects

Start of Block: Block_clinic_visit

UCSF
University of California
San Francisco

While you are taking TB preventive treatment, clinic visits are recommended to make sure you are feeling ok and to help you complete the treatment.
What would make it difficult to come to clinic visits? Click all that apply.

- ▢ Visits are too expensive (e.g., uninsured or high co-payment).
- ▢ It takes a long time to get to the clinic or transportation is too complicated.
- ▢ I want to avoid costs associated with traveling to the clinic.
- ▢ Clinic visits take too much time.
- ▢ I lose income when I go to the clinic.
- ▢ I have to pay for child care or similar costs when I go to the clinic.
- ▢ I have other obligations (e.g., work or school).
- ▢ I have physical limitations that make it difficult for me to travel to the clinic.
- ▢ I have concerns about clinic visits (privacy, do not trust healthcare system, or negative experience).
- ▢ It is not a priority for me, I do not perceive enough benefit from going to the clinic.
- ▢ Other, please specify:________________________________________________

What motivates you to attend clinic visits during TB preventive treatment? Click all that apply.

- ▢ I can discuss any symptoms I may be experiencing with my provider.
- ▢ My provider may have tips and tricks for me to avoid side effects.
- ▢ I can talk to my provider about other health problems.
- ▢ My provider can help me complete preventive treatment.
- ▢ I feel good about clinic visits (e.g., I trust in healthcare or I have prior positive experience).
- ▢ I can get health education and get help so I can manage my own health.
- ▢ My friends and family encourage me to go to the clinic.
- ▢ I can receive incentives or reimbursements when I go to the clinic.
- ▢ Other, please specify:________________________________________________

If your provider recommended clinic visits  during TB preventive treatment, would you accept them and attend?

- ○ No, I would not accept / attend clinic visits.
- ○ Yes, I would accept and attend **one** clinic visit.
- ○ Yes, I would accept and attend **several** clinic visits.
- ○ Maybe, I am not sure.

How would you most prefer to check-in with a provider once a month? Please rank by using drag and drop (1: first choice, 6: least preferred)

______ You talk with your pharmacist on the phone
______ You talk with your pharmacist on video call
______ You visit the pharmacy in-person
______ You talk with your physician on the phone
______ Your talk with your physician on video call
______ You visit your physician in-person

**End of Block: Block_clinic_visit**

---

**Start of Block: Block_blooddraw**

Doctors may use monthly blood tests to check if your liver and kidney are healthy, and if you need to stop TB preventive treatment.

How would you feel about monthly blood tests during TB preventive treatment?

- ○ I would not take preventive treatment if blood tests are required.
- ○ I would accept and take only one blood test before I start treatment.
- ○ I would accept and take monthly blood tests to make sure my liver and kidney are healthy.
- ○ I would take TB preventive treatment but would not accept blood tests.
- ○ I am not sure

**End of Block: Block_blooddraw**

---

**Start of Block: Block_drugdruginteractions**

Most TB preventive treatments can interact with other medications and also with alcohol.

Do you regularly take any of the following? Check all that apply.

- ▢ Drugs prescribed by your doctor
- ▢ Over the counter medications
- ▢ Herbal or other traditional medicine

*Display this choice:If What is your sex? = Female*

- ▢ Contraceptive pills or other hormonal contraceptive

*If Condition: Selected Count Is Equal to 0. Skip To: How often do you have a drink contain....*

If you needed to take TB preventive therapy, would you be willing to stop the medications you indicated above for a few months?

- ○ Yes
- ○ No
- ○ Some of my medications, but not all of them.
- ○ I do not know / do not want to answer

If you needed to take TB preventive therapy, would you be willing to try to find an alternative medication for a few months?

- ○ Yes
- ○ No
- ○ For some of my medications, but not all of them.
- ○ I do not know / do not want to answer

How often do you drink alcohol?

- ○ Never
- ○ Monthly or less
- ○ 2-4 times a month
- ○ 2-3 times a week
- ○ 4 or more times a week

*Display this question: If How often do you drink alcohol? != Never*

During the TB preventive treatment, would you be willing to stop drinking alcohol for a few months?

- ○ Yes
- ○ Maybe
- ○ No
- ○ I do not know / I do not want to answer

**End of Block: Block_drugdruginteractions**

Start of Block: Block_cost

There are some costs related to TB preventive treatment, such as:

- co-payment (the part your insurance does not cover)
- transportation cost (bus ticket, car ride, parking)
- care for relatives or children (e.g., extra child-care while you go to the clinic)

Considering all of the above costs, how much would you be willing to pay for TB preventive treatment in total?

- ○ $0, I would only take the treatment if it was for free
- ○ Up to $50
- ○ Up to $100
- ○ Up to $300
- ○ More than $300

In addition to paying for your treatment, you may also make less money (e.g., you may spend time away from work to attend a clinic visit). How much income will you be willing to lose so you can seek care and prevent **TB disease**?

- ○ $0, I would only take the treatment if I do not lose income
- ○ Up to $50
- ○ Up to $100
- ○ Up to $300
- ○ More than $300

What would you rather be doing instead of seeking care for TB disease prevention? (e.g., leisure activity or work)

________________________________________________________________

How much money is what you described above worth to you (to spend time on your preferred activity instead)? (Select 300 if $300 or more.)

| | 0 30 60 90 120 150 180 210 240 270 300 |
|---|---|
| Worth in $ () | |

End of Block: Block_cost

Start of Block: Block_reinfectionrisk

The TB preventive treatment only treats the current TB germs but does not prevent future infection.

How often do you travel to countries where **TB disease** is common? (e.g., Asia, South America, Central America, Eastern Europe, Africa)

- ○ Every year
- ○ At least once every 2-5 years
- ○ At least once every 6-10 years
- ○ Less often than every 10 years
- ○ I do not anticipate traveling to one of these countries

Among people who travel to countries where **TB disease** is common, about 1 in 100 will get infected (**latent TB**).

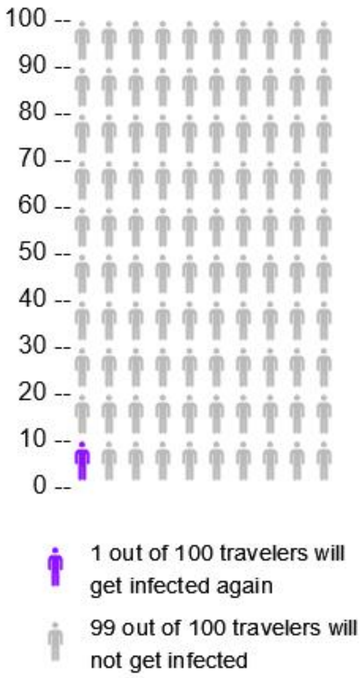


How concerned are you that you will catch TB germs again after completing preventive treatment, when traveling or for other reasons?

- ○ Not at all concerned
- ○ Slightly concerned
- ○ Moderately concerned
- ○ Concerned
- ○ Extremely concerned

Do you have any additional thoughts or comments about how likely or unlikely you might be infected? (Skip if none.)

________________________________________________________________

**End of Block: Block_reinfectionrisk**

**Start of Block: Intro_other**

UCSF University of California San Francisco

We have talked about many aspects of TB preventive treatment. When you consider whether to or not to take TB preventive treatment, which of these matter to you?

| | Not important | Moderately important | Very important |
|---|---|---|---|
| How likely it is for you to develop TB disease when not taking preventive treatment. | ○ | ○ | ○ |
| How likely it is that you will experience nausea or skin rash. | ○ | ○ | ○ |
| How likely it is that you will experience a rare but severe liver damage. | ○ | ○ | ○ |
| Whether there is a need to stop or change at least one of your medications for a few months. | ○ | ○ | ○ |
| Whether there is a need to stop drinking alcohol for a few months. | ○ | ○ | ○ |
| The frequency of clinic visits. | ○ | ○ | ○ |
| The frequency of blood draws. | ○ | ○ | ○ |
| The potential out of pocket costs or loss of income. | ○ | ○ | ○ |
| How likely it is that you will be reinfected with the TB germ. | ○ | ○ | ○ |

Considering TB preventive treatment, are there other factors that are also important to you? (Skip if none.)

________________________________________________________________

**End of Block: Intro_other**

**Start of Block: Block_demographics**

We would like to finish our survey today by understanding a little more about you.
Are you married?

- ○ Married
- ○ Registered partnership
- ○ Single
- ○ Widowed
- ○ Divorced
- ○ Separated

How many persons live with you in your household? (not counting yourself)

- ○ Number of people in household, not including you:________________________________________________

Who lives in your household? Check all that apply.

- ▢ Child or children
- ▢ Senior(s)
- ▢ Persons with condition affecting immune system (such as HIV) or on immune suppressants (such as for auto-immune disease)
- ▢ None of the above

What is your employment status?

- ○ Employed
- ○ Self-employed
- ○ Part-time
- ○ Unemployed or seeking employment
- ○ Student / further education
- ○ Retired
- ○ Other, please specify:________________________________________________
- ○ Prefer not to answer

What is your annual household income (total amount each individual in your household made within the last year)?

- ○ Less than $25'000
- ○ $25'001 to $50'000
- ○ $50'001 to $75'000
- ○ $75'001 to $100'000
- ○ $100'001 to $150'000
- ○ More than $150'000
- ○ Prefer not to answer

Do you have medical insurance?

- ○ Yes
- ○ No
- ○ Other, please specify:________________________________________________

**End of Block: Block_demographics**

**Start of Block: Block_surveyfeedback**

How difficult was it for you to understand the survey questions today?

- ○ Extremely difficult
- ○ Somewhat difficult
- ○ Neither easy nor difficult
- ○ Somewhat easy
- ○ Extremely easy

How difficult was it for you to choose answers to the survey questions today?

- ○ Extremely difficult
- ○ Somewhat difficult
- ○ Neither easy nor difficult
- ○ Somewhat easy
- ○ Extremely easy

*Display this question:*

*If How difficult was it for you to understand the survey questions today? != Extremely easy*

*Or How difficult was it for you to choose answers to the survey questions today? != Extremely easy*

What did you have difficulty understanding or answering?

________________________________________________________________

Would you like to leave any comment about this survey? (Skip if none.)
Click "next" to finish the survey.

________________________________________________________________

**End of Block: Block_surveyfeedback**
