## Supplementary material for "Preferences for treatment for latent tuberculosis infection in primary care among people in the United States at increased risk of tuberculosis: a pilot survey": Online Supplement 2

Online supplemental material 2

### Questionnaire in traditional Chinese

Start of Block: Block_initialCheck

LanguagePreference

我們想邀請您參加調查，以了解您對潛伏性結核感染（LTBI）護理的偏好和觀點。 **潛伏性結核病**患者身體健康，沒有任何症狀，但他們體內攜帶正在休眠的結核病菌。 但這種病菌隨時有繁殖並引致嚴重的活動性結核病的風險。 首先，我們會詢問您一些有關您自己以及您在哪裡接受護理的問題，以確定您是否可以參與本次調查。

在以下語言中，您首選使用哪種語言來完成本次調查？

- ○ 英語 (1)
- ○ 繁體中文 (2)
- ○ 簡體中文 (7)
- ○ 我不習慣用上述任何一種語言完成調查 (3)

---

Age 您的年齡是多少？

- ○ 年： (1) ________________________________________________

---

Sex 您的性別是？

- ○ 男性 (1)
- ○ 女性 (2)
- ○ 其他 (3)
- ○ 不願透露 (4)

---

Education 您完成的最高學歷是什麼？

- ○ 無學歷 (1)
- ○ 幼稚園至 8 年級：小學 / 中學 (2)
- ○ 9 年級或以上但無高中文憑 (5)
- ○ 完成高中或同等學歷，或貿易/技術/職業培訓 (3)
- ○ 高等教育：學院、大學或更高學歷 (4)

---

NEMS_client

您的家庭醫生在東北醫療中心嗎？

- ○ 是 (1)
- ○ 不是，在其它醫療機構 (2)

*Go to Block_ineligible if age below 18, not at NEMS, or not comfortable in any of the offered language.*
*Otherwise continue to Block_CountryOrigin.*

End of Block: Block_initialCheck

Start of Block: Block_ineligible

MessageIneligible 感謝您對本研究調查感興趣並花時間填寫預篩選問題。 根據您的回答，您不符合資格參與本研究調查。 **再次感謝您**的幫助與參與。

*End survey.*

End of Block: Block_ineligible

Start of Block: Block_CountryOrigin

USborn
我們建議出生在**結核病**較常見的國家的人士接受**潛伏性結核病**檢測。 請回答一些關於您出生地以及您的**結核病**或**潛伏性結核病**病史的問題，以檢查您是否符合資格參加本次調查。

您在美國出生嗎？

- 是 (1)
- 不是 (2)

*Skip To: End of Block if Yes*

CountryShortlist 您在哪個國家出生？

- 中國 (1)
- 越南 (2)
- 香港 (3)
- 菲律賓 (4)
- 台灣 (5)
- 墨西哥 (7)
- 印度 (8)
- 泰國 (9)
- 其它 (6)

*Display this question if Other*

CountryOrigin 您在哪個國家出生？

▼ 阿富汗 (1) ... 辛巴威 (1357)

YearImmigration 您什麼時候來到美國的？

- 請輸入年份: YYYY (1) ________________________________________________

- *Go to Block_ineligible if born in any of these countries: San Marino, Saint Kitts and Nevis, Monaco, Barbados, United Arab Emirates, Saint Lucia, United States of America, Slovakia, Israel, Norway, Iceland, Andorra, Grenada, Jamaica, Finland, Hungary, Sweden, Denmark, Czech Republic, Croatia, Greece, Jordan, Slovenia, Netherlands, Cyprus, Switzerland, Ireland, Italy, Antigua and Barbuda, Germany, Austria, Canada, Oman, Luxembourg, United Kingdom of Great Britain and Northern Ireland, Australia, Samoa, New Zealand, Cuba, Tonga, France, Belgium, Spain, Saudi Arabia, Saint Vincent and the Grenadines, Estonia, Lebanon.*
- *Otherwise continue to Block_workexclusions.*

End of Block: Block_CountryOrigin

Start of Block: Block_workexclusions

HealthCareWorker 您是醫護人員嗎?

- ○ 是 (1)
- ○ 否 (2)
- ○ 不知道 (3)

*Skip To: End of Block if Yes*

Congregate
您是否在醫院, 療養院, 監獄, 懲教所或無家可歸者收容所工作嗎?

- ○ 是 (1)
- ○ 否 (2)
- ○ 不知道 (3)

*Go to Block_ineligible if yes for any of these professions.*
*Otherwise continue to Block_TBhistory.*

**End of Block: _workexclusions**

---

**Start of Block: Block_TBhistory**

TBhistory
您曾是否被醫生或公共衛生部門告知患有活動性結核病?
**結核病**是一種肺部疾病。 這是一種嚴重且危及生命的疾病。常見症狀包括咳嗽、發燒、夜間盜汗和體重迅速減輕。如果您患有 **結核病**, 您需要服用 4 種不同的抗生素藥物治療, 治療時間為 6 個月或更長。

- ○ 是 (1)
- ○ 否 (2)
- ○ 不知道 (3)

*Skip To: End of Block if Yes, then go to Block_ineligible. Otherwise continue.*

LTBIexplanation
感染潛伏性結核病的患者 **(潛伏性結核病)** 身體健康並沒有任何症狀，但身體攜帶處於休眠狀態的結核病菌。

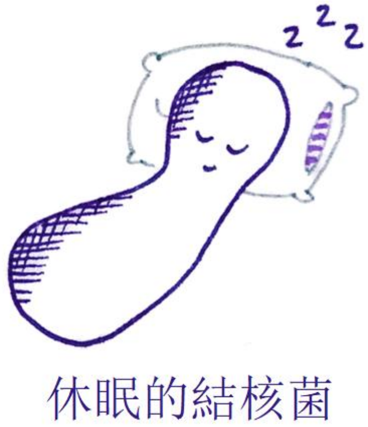

結核菌也能重新繁殖並引致 **結核病**,這是一種嚴重且危及生命的疾病。我們通常建議 **潛伏性結核病** 的患者採取預防性治療，以確保細菌不會重新激活

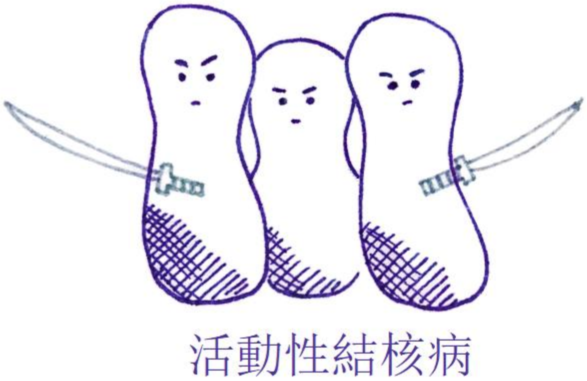

---

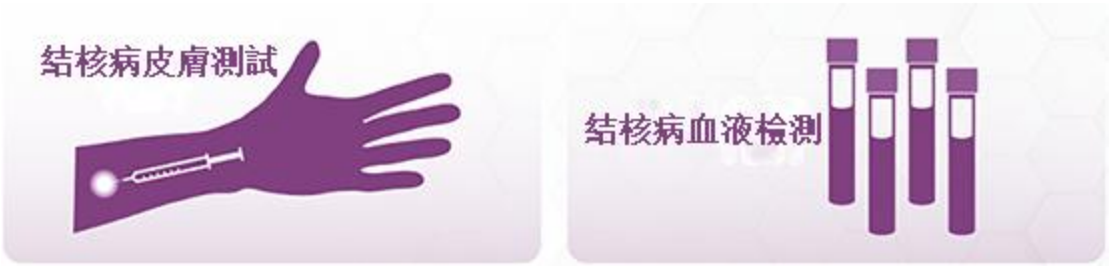

LTBItest 您是否曾經接受過 **潛伏性結核病** 檢測? (皮膚或血液檢測)

- ○ 是 (1)
- ○ 否 (2)
- ○ 不知道 (3)

*Go to Block_iftested if yes.*
*Otherwise skip block and continue to BlockPriorTPT.*

**End of Block: Block_TBhistory**

---

**Start of Block: Block_iftested**

LTBItest_type 您接受了什麼類型的檢測？選出所有適用項。.

- ▢ 結核菌素皮膚檢測 (1)
- ▢ 血液檢測 (2)
- ▢ 不知道 (3)

---

LTBItest_positive 這些檢測結果有否呈陽性（意味著您有結核病菌)?

- ○ 是 (1)
- ○ 否 (2)
- ○ 還沒有收到檢測結果 (3)
- ○ 不知道 (4)

---

*Display this question if Yes*

LTBItest_timing 您 **第一次** 檢測到潛伏性結核病結果 **呈陽性**是什麼時候?

- ○ 一年前或更久之前 (1)
- ○ 不到一年 (2)

**End of Block: Block_iftested**

---

**Start of Block: BlockPriorTPT**

priorTPT 您是否曾接受過 **潛伏性結核病**的預防性治療（健康但感染結核菌）？ 這是一種通過持續數月服用 1-2 種抗生素藥物來降低結核病風險的療法。

- ○ 是 (1)
- ○ 否 (2)
- ○ 不知道 (3)

---

*Display this question if Yes*

timingTPT 您何時開始接受預防性治療?

- ○ 我目前正在接受預防性治療 (1)
- ○ 我這一年內開始治療並已完成或已終止治療。 (2)
- ○ 一年或更久之前 (3)

*Go to Block_ineligible if longer than a year ago.*
*Otherwise continue to BlockDigitalConsent.*

**End of Block: BlockPriorTPT**

---

**Start of Block: BlockDigitalConsent**

ConsentForm

**知情同意書**

本問卷是一項調查研究，您並非必須參加。 如有任何疑問，請聯絡下列研究小組成員。 東北醫療中心與加州大學三藩市分校 (UCSF) 共同參與這項研究。 這項研究領導人為加州大學三藩市山分校的 Priya Shete 博士和東北醫療中心的沈愛梅醫生 (Amy Tang, MD) 。

我們建議非美國出生（即來自大多數其它國家）的人士接受潛伏性結核病篩檢，因此您受邀參與本次研究 調查。 潛伏性結核病患者身體健康，沒有任何症狀，但有發展為活動性結核病（一種嚴重疾病）的風險。

在該研究調查中，研究人員準備了問卷，以更好地了解您對潛伏性結核感染檢測或預防性治療的 看法。 美國疾病管制與預防中心 (CDC) 資助該研究項目，預計約 1400 人將透過不同問卷調查參與研究項目。

**我若參與研究計畫會怎樣？**
如果您同意參與研究項目，您將需要完成問卷並提供回饋。 問卷將透過一系列假設問題詢問您對潛伏性結核感染檢測或預防性治療的看法。 這項問卷調查將幫助我們更了解人們應該何時接受潛伏性結核病檢測和治療。

完成問卷時，請仔細思考並將您的意見或不好理解的問題回饋給我們。 問卷也會有其它額外問題詢問您對於潛伏性結核病的看法。 本問卷調查和訪談大約需要 30 分鐘到 1 小時才能完成。 您可以在平板電腦上完成調查，或請工作人員幫您輸入答案。 現場將有 1-2 名工作人員書面記錄您的回答和回饋。 我們的目標是改善問卷。 您不會被錄音或錄影。 問卷必須由參與者本人在東北醫療中心完成。

**我的資訊會如何使用？**
加州大學三藩市分校的研究人員將使用您的資訊為此研究計畫提供資訊。 研究完成後，我們可能會使用或與其他研究人員分享您的去識別化訊息，用於未來的其它研究。 我們不會透露您的姓名或任何其它個人資訊。 我們不會要求您提供額外的許可證來分享這些去識別化的資訊。

**我或我的隱私會有風險嗎？**
部分問題可能會讓您感到不適或喚起不愉快的回憶。 大多數問題都可以跳過，您也可以隨時終止問卷訪談。 我們會盡全力保護您提供的資訊。 我們不會收集任何可以識別您身分的信息，例如您的姓名或地址。 加州大學的授權代表可能會審查您的研究數據，以監控或管理該研究計畫的進行。

**參與調查對我有好處嗎？**

您不會有直接利益。 雖然問卷調查不能直接幫助您，但可以幫助我們了解人們的檢測和治療偏好，並可能在未來造福其他人。

**我能拒絕參與嗎？**

可以，您並非必須參與本問卷調查。 您的決定絲毫不會影響您在 東北醫療中心的醫療照護服務。

**該問卷調查有金錢獎勵嗎？**

完成本問卷調查後，您將收到由加州大學三藩市分校送出的價值 20 美元的 Target 禮品卡作為酬謝。

**誰可以回答我有關該研究調查的疑問？**
請聯絡沈愛梅醫師 (Amy Tang, MD) 或 Priya Shete 博士。 如果您對自己作為研究參與者的權利有任何問題或疑慮，請致電 415-476-1814 加州大學三藩市分校 機構審查委員會 (UCSF Institutional Review Board)。

Consent_optional
我們需要確保您瞭解此同意書:

您是否可以自願選擇參與本次研究調查？

- ○ 是 (1)
- ○ 否 (2)

Consent_NEMS 您參與本次研究調查會否影響您在東北醫療中心的醫療護理服務？

- ○ 是 (1)
- ○ 否 (2)

Consent_agree
如果您想參與這項研究調查，請選擇“是”，然後點擊 “下一步” 按鈕開始調查。

您是否同意參與這項研究調查？

- ○ 是 (1)
- ○ 否 (2)

*Go to Block_ineligible if no consent.*
*Otherwise continue to StartSurveyC*

**End of Block: BlockDigitalConsent**

---

**Start of Block: StartSurveyC**

interviewer 今天是誰協助您進行本次問卷調查？

- ○ <Community health worker 1> (1)
- ○ <Community health worker 2> (4)
- ○ <Community health worker 3> (10)
- ○ < Researcher 1> (5)
- ○ 沒有人，我自己能完成這項問卷調查。 (8)
- ○ 其他人，請註明： (9) ________________________________________________

---

introtext 這項問卷調查將問及患者決定是否接受**潛伏性結核病**（即身體健康但感染了結核病菌）的預防性治療時可能考慮的不同因素。我们想了解哪些因素對您比較重要。

**End of Block: StartSurveyC**

---

**Start of Block: BlockActiveTB**

Intro_TB_rating **結核病是一種嚴重的疾病。您對結核病帶來的不同影響有多擔心？ 下述每項影響都代表美國每一個 結核病 患者的正常經歷。**

| | 不擔憂 (1) | 中度擔憂 (2) | 非常擔憂 (3) |
|---|---|---|---|
| 住院：您需要住院治療兩週。在醫院裡，您需要被隔離並與他人有極少接觸。 | ○ | ○ | ○ |

| |
|---|
| 健康狀況不佳：經常咳嗽、體重減輕、發燒和盜汗。1 個月後，您發現自己患有結核病。治療兩週後症狀改善，兩個月後症狀消失。 |
| 隔離：出院後，您仍然具有傳染性因此需要留在家中隔離。您至少需要 2 個月後才能回去工作。 |
| 治療（6 個月）：持續 6 個月每天服用藥物；首 2 個月服用 10 種藥物，後 4 個月服用 4 種藥物。您會出現噁心等副作用。您的家庭醫生和護士會每天跟進您的用藥情況。 |
| 對健康的持續影響：許多結核病患者康復後，仍感覺未能回復到之前的那麼健康。 |
| 傳染性：在您未被確診和接受治療之前，您可能會把病菌傳染給與您接觸的親友。 |
| 羞恥: 其他人可能會發現您曾患有結核病。因為您缺席工作、被隔離，並且接觸過您的同事和朋友也會被要求進行結核病感染檢測，所以他們可能會猜測到您患有結核病。 |
| 死亡風險：有些人診斷得太晚。在加州，有 12% 的結核病患者死亡。 |

Intro_TB_top3 您最擔心什麼？請選擇 3 項您擔憂的影響。

*Display each choice if selected as “Very worrisome” or “Moderately worrisome”*

- ▢ 住院：您需要住院治療兩週。在醫院裡，您需要被隔離並與他人有極少接觸。 (1)
- ▢ 健康狀況不佳：經常咳嗽、體重減輕、發燒和盜汗。 1 個月後，您發現自己患有結核病。治療兩週後症狀改善，兩個月後症狀消失。 (2)
- ▢ 隔離：出院後，您仍然具有傳染性因此需要留在家中隔離。您至少需要 2 個月後才能回去工作。 (3)
- ▢ 治療（6 個月）：持續 6 個月每天服用藥物；首 2 個月服用 10 種藥物，後 4 個月服用 4 種藥物。您會出現噁心等副作用。您的家庭醫生和護士會每天跟進您的用藥情況。 (4)
- ▢ 對健康的持續影響：許多結核病患者康復後，仍感覺未能回復到之前的那麼健康。 (5)
- ▢ 傳染性：在您未被確診和接受治療之前，您可能會把病菌傳染給與您接觸的親友。 (6)
- ▢ 羞恥: 其他人可能會發現您曾患有結核病。因為您缺席工作、被隔離，並且接觸過您的同事和朋友也會被要求進行結核病感染檢測，所以他們可能會猜測到您患有結核病。 (7)
- ▢ 死亡風險：有些人診斷得太晚。在加州，有 12% 的結核病患者死亡。 (8)

---

Intro_TB_ranking 您最擔心什麼？請拖放選項來進行排名（1：您最擔憂，3：您最不擔憂）。

*Display each choice if selected among top three*

_______ 住院：您需要住院治療兩週。在醫院裡，您需要被隔離並與他人有極少接觸。 (1)
_______ 健康狀況不佳：經常咳嗽、體重減輕、發燒和盜汗。 1 個月後，您發現自己患有結核病。治療兩週後症狀改善，兩個月後症狀消失。 (2)
_______ 隔離：出院後，您仍然具有傳染性因此需要留在家中隔離。您至少需要 2 個月後才能回去工作。 (3)
_______ 治療（6 個月）：持續 6 個月每天服用藥物；首 2 個月服用 10 種藥物，後 4 個月服用 4 種藥物。您會出現噁心等副作用。您的家庭醫生和護士會每天跟進您的用藥情況。 (4)
_______ 對健康的持續影響：許多結核病患者康復後，仍感覺未能回復到之前的那麼健康。 (5)
_______ 傳染性：在您未被確診和接受治療之前，您可能會把病菌傳染給與您接觸的親友。 (6)
_______ 羞恥: 其他人可能會發現您曾患有結核病。因為您缺席工作、被隔離，並且接觸過您的同事和朋友也會被要求進行結核病感染檢測，所以他們可能會猜測到您患有結核病。 (7)
_______ 死亡風險：有些人診斷得太晚。在加州，有 12% 的結核病患者死亡。 (8)

Page Break

Intro_LTBI_TPT
**潛伏性結核病**患者 (身體健康但感染結核病菌) 可能會也可能不會引致 **結核病**。一個人罹患**結核病** 的可能性取決於其年齡、吸煙狀況、整體健康狀況以及其它因素。美国疾病控制与预防中心 (CDC) 建議進行結核病預防性治療。
但是，有些人會在預防結核病和 3 至 4 個月的治療療程間進行權衡 。更有可能患上**結核病**的人可能會從結核病預防性治療中獲益更多。對於不太可能患上**結核病**的人，即使接受結核病預防性治療也收益不大，僅能進一步降低罹患結核病的可能性。我們想要了解您在患**結核病**的風險有多大的情況下才會採取預防性治療。

對於下一個問題，請假設您患有**潛伏性結核病**。在未來 10 年內您有罹患**結核病**的風險。

*Randomize to BlockThreshold5 or BlockThreshold10, i.e., starting with 5% or 10% risk of TB disease. Only show one of the two blocks.*

**End of Block: BlockActiveTB**

**Start of Block: BlockThreshold5**

0.05 如果像您一樣的 100 人中有 5 個人**在未來 10 年內會罹患結核病**，您是否願意接受治療？

- ○ 是，我願意接受預防性治療 (1)
- ○ 不，我不願意接受治療 (2)

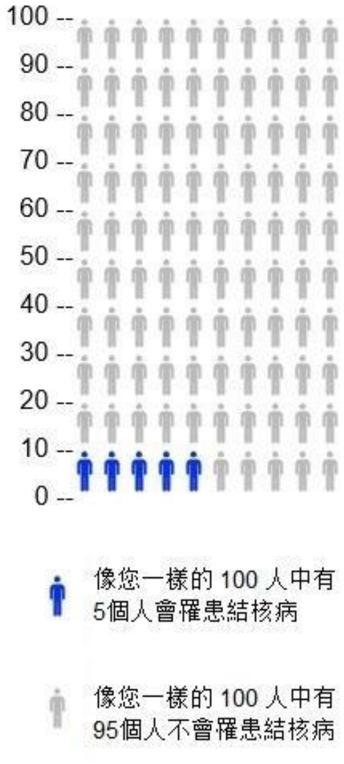

*If yes, skip to “less than 1 out of 100”. If no, continue to 50%. Follow the logic shown below. Arrows to the left mean “yes”, arrows to the right mean “no”. A straight arrow down to “stop” means go to the End of Block, regardless of the answer.*

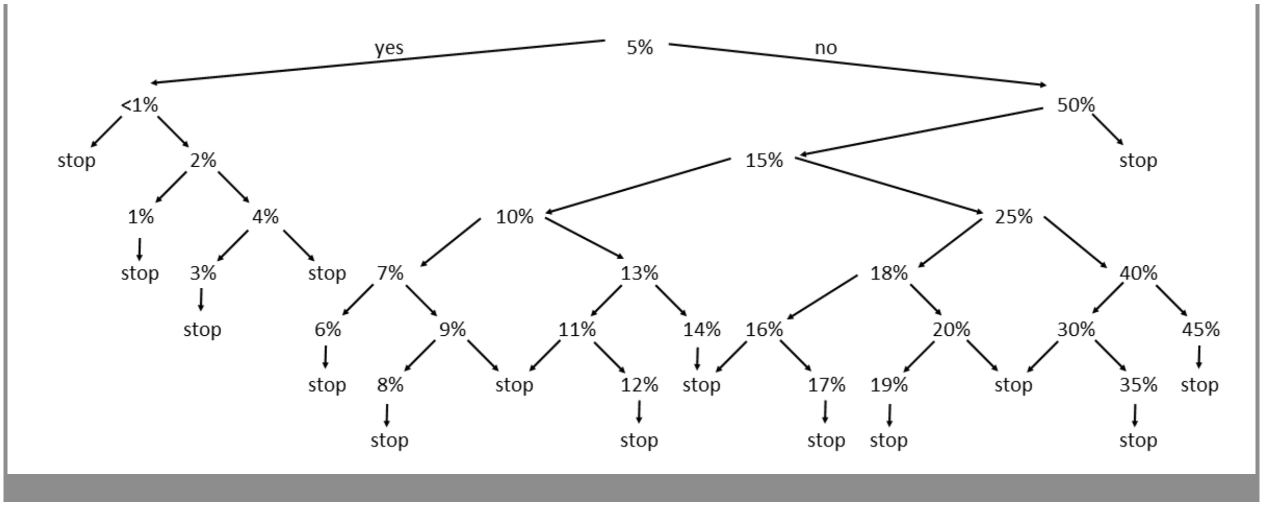

*At the end of this block, skip to Block_TBrisk_sliders.*

End of Block: BlockThreshold5

---

Start of Block: BlockThreshold10

T0.10 如果像您一樣的 100 人中有 10 個人**在未來 10 年內會罹患結核病**，您是否願意接受治療？

- 是，我願意接受預防性治療 (1)
- 不，我不願意接受治療 (2)

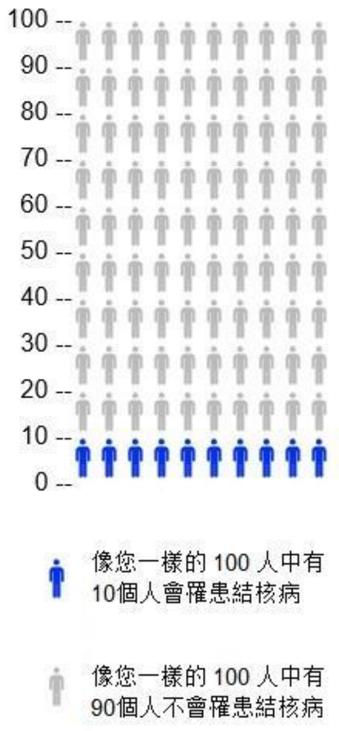

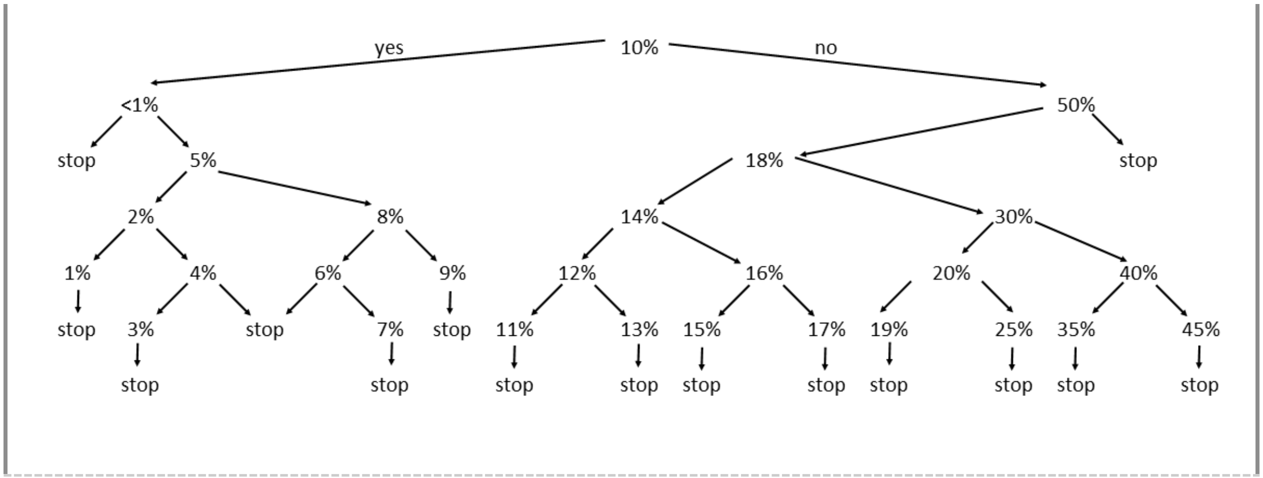

[We acknowledge that there was a translation error in the legend of the icon array for the risk below 1%. However, the question was translated and administered correctly, and we are confident the question was interpreted correctly by all participants.] Here is the figure legend with the error:]

T0.001 如果像您一樣的 100 人中有少於 1 個人**在未來 10 年內會患上結核病**，您是否願意接受治療？

- 是，我願意接受預防性治療 (1)
- 不，我不願意接受治療 (2)

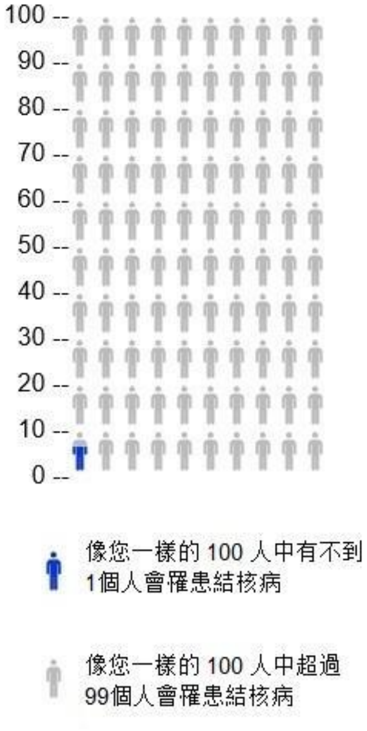

*At the end of this block, skip to Block_TBrisk_sliders.*

**End of Block: BlockThreshold10**

Start of Block: Block_TBrisk_sliders

TBsliders_intro 我們現在將討論時間如何影響您願意接受結核病預防治療的想法。
[randomized order of questions: 10 years, 2 years, life time risk]

TBslider_10y
考慮一下您**在未來 10 年內罹患結核病**的機率。
要多高的患病機率，您才會考慮接受預防性治療?（例如，想哪一種可能性較小?） 點擊選擇合適的答案。

| | 低機率(%) | | | | | | 高機率(%) | | | | |
|---|---|---|---|---|---|---|---|---|---|---|---|
| | 0 | 10 | 20 | 30 | 40 | 50 | 60 | 70 | 80 | 90 | 100 |
| **未來 10 年**罹患結核病的機率 (%) () | | | | | | | | | | | |

TBslider_2y
假設您在**未來 2 年內**可能會罹患**結核病**。
要多高的患病機率，您才會考慮接受預防性治療？

| | 低機率(%) | | | | | | 高機率(%) | | | | |
|---|---|---|---|---|---|---|---|---|---|---|---|
| | 0 | 10 | 20 | 30 | 40 | 50 | 60 | 70 | 80 | 90 | 100 |
| **未來 2 年**罹患結核病的機率 (%) () | | | | | | | | | | | |

TBslider_lifetime 考慮一下您**終生患有結核病**的機率。 例如，假設您活到 90 歲，請考慮您在 90 歲之前患有**結核病**的風險。 罹患**結核病**的機率要多高，才會接受預防性治療？

| | 低機率(%) | | | | | | 高機率(%) | | | | |
|---|---|---|---|---|---|---|---|---|---|---|---|
| | 0 | 10 | 20 | 30 | 40 | 50 | 60 | 70 | 80 | 90 | 100 |
| **終生**罹患結核病的機率 (%) () | | | | | | | | | | | |

End of Block: Block_TBrisk_sliders

Start of Block: Block_TBrisk_sliders2

TBrisk_choice_time 您的醫師向您解釋罹患**結核病**的機率時，考慮到您自身罹患結核病的風險的情況下，什麼時間範圍對您來說最重要？

- ○ 1 年 (6)
- ○ 2 年 (1)
- ○ 5 年 (2)
- ○ 10 年 (3)
- ○ 15 年 (7)
- ○ 20 年 (8)
- ○ 50 年 (9)
- ○ 終身風險 (4)
- ○ 其它，請註明： (5) ________________________________________________

End of Block: Block_TBrisk_sliders2

---

Start of Block: Side effects

Intro_AEs
結核病預防性治療有時可能會引起副作用，例如噁心或持續約一週的皮疹。 這些副作用會自行消失。
多數患者無需看醫生。

---

accept_AE 即使可能出現暫時的噁心或皮疹，您也會接受結核病預防性治療嗎？

- ○ 是 (1)
- ○ 可能 (2)
- ○ 否 (3)

---

accept_AE_0.3 每個人出現副作用（如噁心或持續約一週的皮疹）的機率是不同的。 假設每 100 個像您一樣的人中有 30 人出現噁心或皮疹。 您還願意接受結核病預防性治療嗎？

- ○ 是 (1)
- ○ 可能 (2)
- ○ 否 (3)

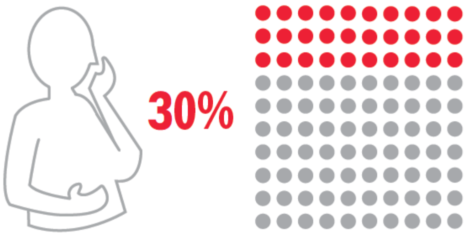

---

accept_AE_1.0 假設所有人都會感到噁心或持續約一週的皮疹。 您還願意接受結核病預防性治療嗎？

- 是 (1)
- 可能 (2)
- 否 (3)

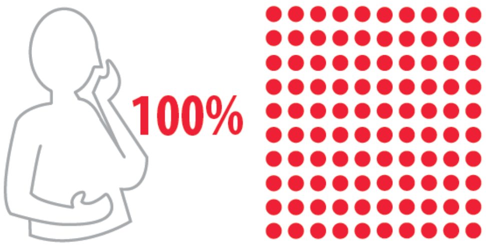

Page Break

Intro_Hepatotoxicity 結核病預防性治療也可能導致嚴重的副作用，例如肝功能損傷。 每 100 人中有 0 到 5 人會出現肝功能損傷，出現這種副作用的人需要去醫院就診。

accept_AE_hepato 鑑於這種肝功能損傷機率（每 100 人中有 5 人）， 您還願意接受結核病預防性治療嗎？

- 是 (1)
- 可能 (2)
- 否 (3)

value_AE
您更擔心的是： 比較不常見的肝功能損傷（每 100 人中有 5 人會出現）或 比較常見的噁心或持續一週的皮疹（100 人中有 30 人會出現）？

- 我更擔心肝功能損傷 (1)
- 我更擔心噁心或皮疹 (2)
- 兩者我都擔心 (3)
- 兩者我都不擔心，預防結核病更重要。 (4)
- 其它，請註明： (5) ________________________________________________

accept_AE_open_2 針對結核病預防性治療的副作用，您有什麼意見？ （沒有則跳過此問題）

________________________________________________________________

End of Block: Side effects

Start of Block: Block_clinic_visit

UCSF University of California San Francisco

reason_against_visit
在接受結核病預防性治療期間，我們建議您前往診所就診，以確保您感覺良好並幫助您完成治療。
哪些因素會導致去診所就診變得困難？ 選擇所有適用的選項。

- ▢ 就診費用太昂貴（例如，沒有保險或高額共付額）。 (10)
- ▢ 去診所路途太長或交通過於複雜。 (8)
- ▢ 我想避免因前往診所而產生的費用。 (1)
- ▢ 去診所太花時間。 (2)
- ▢ 我去診所時就會失去收入。 (4)
- ▢ 我去診所就必須支付託兒費或類似費用。 (5)
- ▢ 我有其它要事（如工作或上學）。 (13)
- ▢ 我的身體狀況受限，很難前往診所。 (14)
- ▢ 我對去診所就診有顧慮（隱私問題、不信任醫療保健系統或負面的就診經驗）。 (15)
- ▢ 這對我來說不是優先考慮的事情，去診所也不會有多大幫助。 (16)
- ▢ 其它，請註明： (9) ________________________________________________

---

reason_for_visits 在接受預防性治療期間，促使您去診所就診的動力是什麼？ 選擇所有適用的選項。

- ▢ 我可以與家庭醫生討論可能出現的症狀。 (6)
- ▢ 我的家庭醫生可能有辦法幫我避免副作用。 (12)
- ▢ 我可以與家庭醫生討論我的其它健康問題。 (5)
- ▢ 我的家庭醫生能幫助我完成預防性治療。 (4)
- ▢ 我對去診所就診感覺良好（例如：我信任醫療系統，或之前有良好的就診經驗）。 (13)
- ▢ 我可以獲得幫助及健康知識，方便我管理自己的健康狀況。 (14)
- ▢ 我的親友建議我到診所就診。 (15)
- ▢ 去診所就診可以獲得獎勵或報銷。 (16)
- ▢ 其它，請註明： (9) ________________________________________________

---

barrier_clinicvisit 如果您的家庭醫生建議您在接受結核病預防性治療期間到診所就診，您會接受並到診所就診嗎？

- ○ 不，我不會接受或到診所就診。 (1)
- ○ 是，我願意接受並到診所進行**一次**就診。 (2)
- ○ 是，我願意接受並到診所進行**多次**就診。 (4)
- ○ 可能，我不確定。 (3)

---

ranking_visittype 您最希望透過何種方式與您的家庭醫師進行每月一次的溝通？ 請拖放選項進行排名（1：首選，6：最後選擇）

______ 與您的藥師電話溝通 (1)
______ 與您的藥劑師視訊溝通 (6)
______ 親自前往藥房 (2)
______ 與您的家庭醫生電話溝通 (3)
______ 與您的家庭醫生視訊溝通 (4)
______ 本人就診 (5)

End of Block: Block_clinic_visit

Start of Block: Block_blooddraw

UCSF
University of California
San Francisco

Intro_blooddraw 醫生可能會要求您每月抽血檢查，以此監測您的肝腎是否健康，並評估您是否需要停止結核病預防性治療。

---

barrier_bloodtests 您對於結核病預防性治療期間的每月抽血檢查有何看法？

- ○ 如果必須抽血檢查，我不會接受預防性治療。 (1)
- ○ 我只會在開始預防性治療之前做一次抽血檢查。 (2)
- ○ 我願意接受每月一次的抽血檢查，以確保我的肝腎處於健康狀態。 (4)
- ○ 我願意接受結核病預防性治療，但不願接受抽血檢查。 (5)
- ○ 我不確定 (3)

End of Block: Block_blooddraw

---

Start of Block: Block_drugdruginteractions

Intro_ddi 大多數結核病預防性治療會與其它藥物及酒精會產生交互作用。

---

medications 您經常服用以下任何一種藥物嗎？ 勾選所有適用的項目。

- ▢ 您家庭醫師開的藥 (1)
- ▢ 非處方藥 (2)
- ▢ 草藥或其它傳統藥物 (3)

*Display this choice:If What is your sex? = Female*

- ▢ 避孕藥或其他荷爾蒙避孕藥 (4)

*If Condition: Selected Count Is Equal to 0. Skip To: How often do you have a drink contain....*

---

stop_medications 假設您需要接受結核病預防性治療，您願意停用上述藥物幾個月嗎？

- ○ 是 (1)
- ○ 否 (2)
- ○ 可以停用一些藥，但不能全停。 (4)
- ○ 我不知道/不想回答 (3)

---

replace_medications 假設您需要接受結核病預防性治療，您願意嘗試服用其它替代藥物幾個月嗎？

- ○ 是 (1)
- ○ 否 (2)
- ○ 可以替換一些藥，但不能全換。 (4)
- ○ 我不知道/不想回答 (3)

---

freq_alcohol 您喝酒的頻率如何？

- ○ 從來不喝 (1)
- ○ 每個月喝一次或更少 (2)
- ○ 每月 2-4 次 (3)
- ○ 每週 2-3 次 (4)
- ○ 每週 4 次或更多 (5)

---

Display this question: If How often do you drink alcohol? != Never

stop_alcohol 假設您需要接受結核病預防性治療，您願意停酒幾個月嗎？

- ○ 是 (1)
- ○ 可能 (2)
- ○ 否 (3)
- ○ 我不知道/不想回答 (4)

End of Block: Block_drugdruginteractions

Start of Block: Block_cost

Intro_cost

結核病預防性治療可能產生一些費用，例如：

共付額（您的保險不承保的部分）

交通費用（巴士票、計程車、停車費）

照顧親屬或孩子的費用（例如，去診所時需要找其他人看護兒童）

threshold_cost 綜合考慮上述所有費用，您願意支付總額多少錢接受結核病預防性治療？

- ○ $0，我只接受免費治療 (1)
- ○ 最多$50 (2)
- ○ 最多$100 (3)
- ○ 最多$300 (4)
- ○ 大於$300 (5)

lost_income 除了額外支付治療費用，您的收入也可能會減少（例如，您可能需要請假才能去就診）。 您願意承擔因接受**結核病**預防性治療導致的收入損失是多少？

- ○ $0，我只會在不影響收入的情況下接受治療 (1)
- ○ 最多$50 (2)
- ○ 最多$100 (3)
- ○ 最多$300 (4)
- ○ 大於$300 (5)

opportunity_cost 與尋求結核病預防性護理相比，您更願意做什麼？ （例如休閒活動或工作）

________________________________________________________________

opportunity_quant

您願意為上述活動花多少錢（將時間花在您喜歡的活動上）？

（如果大於等於$300，請選擇 $300。）

| | 0 30 60 90 120 150 180 210 240 270 300 |
|---|---|
| 以美元計算 () | |

End of Block: Block_cost

Start of Block: Block_risk

Intro_reinfection
結核病的預防性治療只能治療目前的結核病菌，但不能預防未來的感染。

reinf_travel 您前往**結核病**常見國家（例如亞洲、南美洲、中美洲、東歐、非洲）的頻率是多少？

- ○ 每年都去 (1)
- ○ 每 2-5 年去一次 (2)
- ○ 每 6-10 年去一次 (3)
- ○ 每 10 年或更久才去一次 (4)
- ○ 我不打算前往這些國家 (5)

reinf_riskperceived
前往**結核病**常見國家旅行的人當中，大約每 100 人中有 1 人會感染（**潛伏性結核病**）。

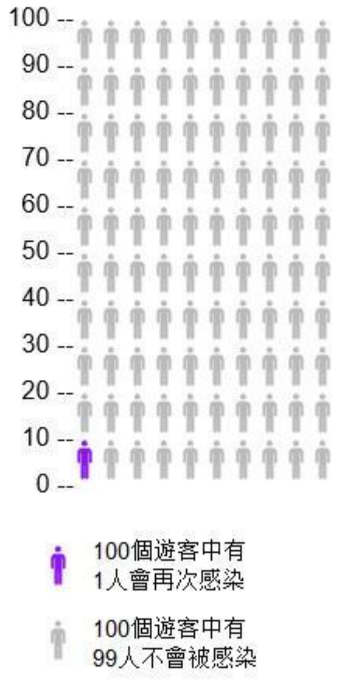

您會否擔心完成預防性治療後，會因旅行或其它原因再次感染結核病菌？

- ○ 一點也不擔心 (1)
- ○ 有一點擔心 (2)
- ○ 一般擔心 (3)
- ○ 擔心 (4)
- ○ 非常擔心 (5)

reinf_open 您對於被感染的可能性是否有其它意見或看法？（沒有則跳過）

________________________________________________________________

End of Block: Block_reinfectionrisk

Start of Block: Intro_other

Overall_rating 我們已經討論了結核病預防性治療的許多方面。 當您在考慮是否接受結核病預防性治療時，下列哪一項對您來說很重要？

| | 不重要 (1) | 一般重要 (2) | 非常重要 (3) |
|---|---|---|---|
| 如果不接受預防性治療，您罹患結核病的可能性有多大。 (1) | ○ | ○ | ○ |
| 您出現噁心或皮疹的可能性有多高。 (2) | ○ | ○ | ○ |
| 您出現罕見但嚴重的肝損傷的可能性有多大。 (3) | ○ | ○ | ○ |
| 是否需要換藥或停藥幾個月。 (4) | ○ | ○ | ○ |
| 是否需要停止飲酒幾個月。 (5) | ○ | ○ | ○ |
| 去診所就診的頻率。 (6) | ○ | ○ | ○ |
| 抽血檢查的頻率。 (7) | ○ | ○ | ○ |
| 可能產生的自付費用或收入損失。 (8) | ○ | ○ | ○ |
| 您再次感染結核菌的可能性有多高。 (9) | ○ | ○ | ○ |

open_other 考慮結核病預防性治療時，還有其它對您來說也很重要的因素嗎？ （如果沒有則跳過。）

________________________________

End of Block: Intro_other

Start of Block: Block_demographics

civilstatus

完成今天的問卷調查之前，我們想要更了解您。

您結婚了嗎？

- ○ 已婚 (1)
- ○ 同居伴侶 (2)
- ○ 單身 (3)
- ○ 喪偶 (4)
- ○ 離婚 (5)
- ○ 分居 (6)

---

householdsize 不包含您本人，您家中有多少人與您同住？

- ○ 家庭成員人數（不包含您本人）： (1) ________________________________________________

---

householdtype 哪些家庭成員住在您家？ 勾選所有適用項。

- ▢ 子女（們） (1)
- ▢ 老人（們) (2)
- ▢ 患有影響免疫系統疾病（例如愛滋病毒）或正在服用免疫抑制劑（例如自體免疫疾病）的人 (3)
- ▢ 以上全都沒有 (4)

---

workstatus 您的就業狀況是

- ○ 受僱 (1)
- ○ 自僱 (2)
- ○ 兼職 (3)
- ○ 未就業或正在找工作 (4)
- ○ 學生/延續教育 (5)
- ○ 退休 (6)
- ○ 其它，請註明： (7) ________________________________________________
- ○ 不願回答 (8)

---

income 您的家庭年收入是多少（您家庭中每個人去年的收入總和）？

- ○ 低於$25'000 (1)
- ○ $25'001 至$50'000 (2)
- ○ $50'001 至$75'000 (3)
- ○ $75'001 至$100'000 (4)
- ○ $100'001 至$150'000 (5)
- ○ 高於$150'000 (6)
- ○ 不願回答 (7)

---

healthinsurance 您有醫療保險嗎？

- ○ 是 (1)
- ○ 否 (2)
- ○ 其它，請註明： (3) ________________________________________________

End of Block: Block_demographics

---

Start of Block: Block_surveyfeedback

UCSF University of California San Francisco

diff_understand 您認為該問卷調查的問題很難理解嗎？

- 很難 (9)
- 比較困難 (10)
- 一般 (11)
- 比較容易 (12)
- 非常容易 (13)

---

diff_choose 您今天選擇該問卷調查問題的答案時有多困難？

- 很難 (11)
- 比較困難 (12)
- 一般 (13)
- 比較容易 (14)
- 非常容易 (15)

---

*Display this question:*

*If How difficult was it for you to understand the survey questions today? != Extremely easy*

*Or How difficult was it for you to choose answers to the survey questions today? != Extremely easy*

diff_open 您在理解或回答問題時遇到什麼困難？

________________________________________________________________

---

survey_open

關於此次調查，您有何意見及建議嗎？（沒有則跳過。）

點擊「下一步」完成調查。

________________________________________________________________

**End of Block: Block_surveyfeedback**
