## Supplementary material for "Preferences for treatment for latent tuberculosis infection in primary care among people in the United States at increased risk of tuberculosis: a pilot survey": Online Supplement 3

Online supplemental material 3

### Questionnaire in simplified Chinese

Start of Block: Block_initialCheck

LanguagePreference
我们想邀请您参加一项调查，以了解您对潜伏性结核感染（LTBI）护理的偏好和观点。**潜伏性结核病**患者身体健康，没有任何症状，但他们体内携带正在休眠的结核病菌。但这种病菌随时有繁殖并引致严重的活动性结核病的风险。首先，我们会询问您一些有关您自己以及您在哪里接受护理的问题，以确定您是否可以参与本次调查。
在以下语言中，您首选使用哪种语言来完成本次调查？

- 英语 (1)
- 繁体中文 (2)
- 简体中文 (7)
- 我不习惯用上述任何一种语言完成调查 (3)

---

Age 您的年龄是多少？

- 年： (1) ________________________________________________

---

Sex 您的性别是什么？

- 男性 (1)
- 女性 (2)
- 其他 (3)
- 不愿透露 (4)

---

Education 您完成的最高学历是什么？

- 无学历 (1)
- 幼儿园至 8 年级：小学 / 中学 (2)
- 9 年级或以上但无高中文凭 (5)
- 完成高中或同等学历，或贸易/技术/职业培训 (3)
- 高等教育：学院、大学或更高学历 (4)

---

NEMS_client
您的家庭醫生在東北醫療中心吗？

- 是 (1)
- 不是，在其它医疗机构 (2)

*Go to Block_ineligible if age below 18, not at NEMS, or not comfortable in any of the offered language.*
*Otherwise continue to Block_CountryOrigin.*

End of Block: Block_initialCheck

Start of Block: Block_ineligible

MessageIneligible 感谢您对本研究调查感兴趣并花时间填写预筛选问题。 根据您的回答，您不符合条件参与这项研究调查。 **再次感谢您**的帮助和参与。

*End survey.*

End of Block: Block_ineligible

---

Start of Block: Block_CountryOrigin

USborn
我们建议出生在**结核病**较常见的国家的人士接受**潜伏性结核病**检测。 请回答一些关于您出生地以及您的**结核病**或**潜伏性结核病**病史的问题，以检查您是否符合条件参加本次调查。

您在美国出生吗？

- ○ 是 (1)
- ○ 不是 (2)

*Skip To: End of Block if Yes*

CountryShortlist 您在哪个国家出生？

- ○ 中国 (1)
- ○ 越南 (2)
- ○ 香港 (3)
- ○ 菲律宾 (4)
- ○ 台湾 (5)
- ○ 墨西哥 (7)
- ○ 印度 (8)
- ○ 泰国 (9)
- ○ 其它 (6)

*Display this question if Other*

CountryOrigin 您在哪个国家出生？

▼ 阿富汗 (1) ... 津巴布韦 (1357)

YearImmigration 您什么时候來到美国的？

- ○ 请输入年份: YYYY (1) ________________________________________________

*Go to Block_ineligible if born in any of these countries: San Marino, Saint Kitts and Nevis, Monaco, Barbados, United Arab Emirates, Saint Lucia, United States of America, Slovakia, Israel, Norway, Iceland, Andorra, Grenada, Jamaica, Finland, Hungary, Sweden, Denmark, Czech Republic, Croatia, Greece, Jordan, Slovenia, Netherlands, Cyprus, Switzerland, Ireland, Italy, Antigua and Barbuda, Germany, Austria, Canada, Oman, Luxembourg, United Kingdom of Great Britain and Northern Ireland, Australia, Samoa, New Zealand, Cuba, Tonga, France, Belgium, Spain, Saudi Arabia, Saint Vincent and the Grenadines, Estonia, Lebanon.*

*Otherwise continue to Block_workexclusions.*

End of Block: Block_CountryOrigin

---

Start of Block: Block_workexclusions

HealthCareWorker 您是医护人员吗?

- 是 (1)
- 否 (2)
- 不知道 (3)

*Skip To: End of Block if Yes*

Congregate
您是否在医院, 疗养院, 监狱, 惩教所或无家可归者收容所工作吗?

- 是 (1)
- 否 (2)
- 不知道 (3)

*Go to Block_ineligible if yes for any of these professions.*
*Otherwise continue to Block_TBhistory.*

**End of Block: Block_workexclusions**

---

**Start of Block: Block_**

TBhistory
您曾是否被医生或公共卫生部门告知患有活动性结核病?

**结核病**是一种肺部疾病。这是一种严重且危及生命的疾病。常见症状包括咳嗽、发烧、夜间盗汗和体重迅速减轻。如果您患有 **结核病**, 您需要服用 4 种不同的抗生素药物治疗, 治疗时间为 6 个月或更长。

- 是 (1)
- 否 (2)
- 不知道 (3)

*Skip To: End of Block if Yes, then go to Block_ineligible. Otherwise continue.*

LTBIexplanation
感染潜伏性结核病的患者 (**潜伏性结核病**) 身体健康并没有任何症状，但身体携带处于休眠状态的结核病菌。

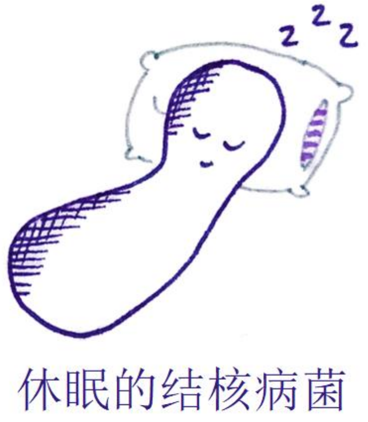


结核菌也能重新繁殖并引致**结核病**,这是一种严重且危及生命的疾病。我们通常建议**潜伏性结核病** 的患者采取预防性治疗，以确保细菌不会重新激活

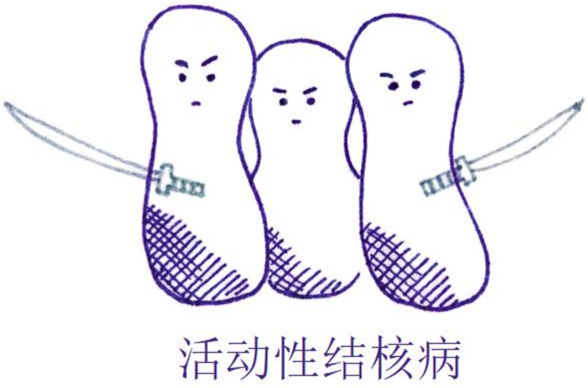


---

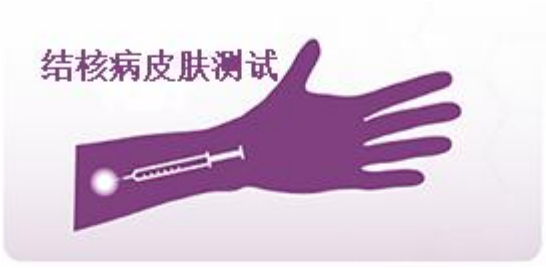


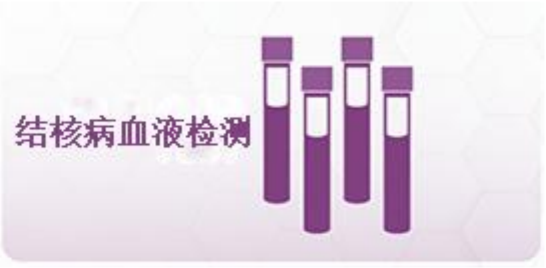


LTBItest 您是否曾经接受过 **潜伏性结核病** 检测? (皮肤或血液检测)

- ○ 是 (1)
- ○ 否 (2)
- ○ 不知道 (3)

*Go to Block_iftested if yes.*
*Otherwise skip block and continue to BlockPriorTPT.*

**End of Block: Block_TBhistory**

---

**Start of Block: Block_iftested**

LTBItest_type 您接受了什么类型的检测？选出所有适用项。 .

- ▢ 结核菌素皮肤检测 (1)
- ▢ 血液检测 (2)
- ▢ 不知道 (3)

---

LTBItest_positive 这些检测结果有否呈阳性（意味着您有结核病菌?

- ○ 是 (1)
- ○ 否 (2)
- ○ 还没有收到检测结果 (3)
- ○ 不知道 (4)

---

*Display this question if Yes*

LTBItest_timing 您 **第一次** 检测到潜伏性结核病结果 **呈阳性**是什么时候?

- ○ 一年前或更久之前 (1)
- ○ 不到一年 (2)

**End of Block: Block_iftested**

---

**Start of Block: BlockPriorTPT**

priorTPT 您是否曾接受过**潜伏性结核病**的预防性治疗（健康但感染结核菌）？ 这是一种通过持续数月服用 1-2 种抗生素药物来降低结核病风险的疗法。

- ○ 是 (1)
- ○ 否 (2)
- ○ 不知道 (3)

---

*Display this question if Yes*

timingTPT 您何时开始接受预防性治疗?

- ○ 我目前正在接受预防性治疗 (1)
- ○ 我这一年内开始治疗并已完成或已终止治疗。 (2)
- ○ 一年或更久之前 (3)

*Go to Block_ineligible if longer than a year ago.*
*Otherwise continue to BlockDigitalConsent.*

**End of Block: BlockPriorTPT**

---

**Start of Block: BlockDigitalConsent**

ConsentForm
**知情同意书**
本问卷是一项调查研究，您并非必须参加。如有任何疑问，请联系下列研究小组成员。 东北医疗中心与加州大学三藩市分校 (UCSF) 共同参与这项研究。该项研究领导人为加州大学三藩市山分校的 Priya Shete 博士和东北医疗中心的沈

爱梅医生 (Amy Tang, MD) 。

我们建议非美国出生（即来自大多数其它国家）的人士接受潜伏性结核病筛查，因此您受邀参与本次研究调查。潜伏性结核病患者身体健康，没有任何症状，但存在发展为活动性结核病（一种严重疾病）的风险。

在该研究调查中，研究人员准备了调查问卷，以更好地了解您对潜伏性结核感染检测或预防性治疗的看法。美国疾病控制与预防中心 (CDC) 资助该研究项目，预计约 1400 人将通过不同问卷调查参与该研究项目。

**我若参与该研究项目会怎样？**
如果您同意参与该研究项目，您将需要完成调查问卷并提供反馈。该问卷将通过一系列假设问题询问您对潜伏性结核感染检测或预防性治疗的看法。该问卷调查将帮助我们更好地了解人们应该何时接受潜伏性结核病检测和治疗。

完成问卷时，请仔细思考并将您的意见或不好理解的问题反馈给我们。问卷也会有其它额外问题询问您对于潜伏性结核病的看法。本次问卷调查和采访大约需要 30 分钟到 1 小时才能完成。您可以在平板电脑上完成调查，或请工作人员帮助您输入答案。现场将有 1-2 名工作人员书面记录您的回答和反馈。我们的目标是改善调查问卷。您不会被录音或录像。问卷必须由参与者本人在东北医疗中心完成。

**我的信息会被如何使用？**
加州大学三藩市分校的研究人员将使用您的信息为该研究项目提供信息。该研究完成后，我们可能会使用或与其他研究人员共享您的去识别化信息，用于未来的其它研究。我们不会透露您的姓名或任何其它个人信息。我们不会要求您提供额外的许可来共享这些去识别化的信息。

**我或我的隐私会有风险吗？**
部分问题可能会让您感到不适或唤起不愉快的回忆。大多数问题都可以跳过，您也可以随时终止问卷采访。我们会尽全力保护您提供的信息。我们不会收集任何可以识别您身份的信息，例如您的姓名或地址。加州大学的授权代表可能会审查您的研究数据，以监控或管理该研究项目的进行。

**参与调查对我有好处吗？**
您不会有直接利益。虽然问卷调查不能直接帮助您，但可以帮助我们了解人们的检测和治疗偏好，并可能在未来造福其他人。

**我能拒绝参与吗？**
可以，您并非必须参与本次问卷调查。您的决定丝毫不会影响您在 东北医疗中心的医疗护理服务。

**该问卷调查有金钱奖励吗？**
完成本次问卷调查后，您将收到由加州大学三藩市分校送出的价值 20 美元的 Target 礼品卡作为酬谢。

**谁可以回答我有关该研究调查的疑问？**
请联系沈爱梅医生 (Amy Tang, MD) 或 Priya Shete 博士。如果您对自己作为研究参与者的权利有任何问题或疑虑，请致电 415-476-1814 加州大学三藩市分校 机构审查委员会 (UCSF Institutional Review Board)。

Consent_optional
我们需要确保您了解此同意书:

您是否可以自愿选择参与本次研究调查？

- 是 (1)
- 否 (2)

Consent_NEMS 您参与本次研究调查会否影响您在东北医疗中心的医疗护理服务？

- 是 (1)
- 否 (2)

Consent_agree
如果您想参与这项研究调查，请选择“是”，然后点击 “下一步” 按钮开始调查。

您是否同意参与这项研究调查？

- 是 (1)
- 否 (2)

*Go to Block_ineligible if no consent.*
*Otherwise continue to StartSurveyC*

**End of Block: BlockDigitalConsent**

---

**Start of Block: StartSurveyC**

interviewer 今天是谁协助您进行本次问卷调查？

- <Community health worker 1> (1)
- <Community health worker 2> (4)
- <Community health worker 3> (10)
- < Researcher 1> (5)
- 没有人，我自己能完成这项问卷调查。 (8)
- 其他人，请注明： (9) ________________________________________________

---

introtext 这项问卷调查将问及患者决定是否接受**潜伏性结核病**（即身体健康但感染了结核病菌）的预防性治疗时可能考虑的不同因素。我们想了解哪些因素对您比较重要。

**End of Block: StartSurveyC**

---

**Start of Block: BlockActiveTB**

Intro_TB_rating **结核病是一种**严重的疾病。您对**结核病**带来的不同影响有多担心? **下述每**项影响都代表美国每一个 **结核病 患者的正常**经历。

| | 不担忧 (1) | 中度担忧 (2) | 非常担忧 (3) |
|---|---|---|---|
| 住院：您需要住院治疗两周。在医院里，您需要被隔离并与他人有极少接触。 | ○ | ○ | ○ |
| 健康状况不佳：经常咳嗽、体重减轻、发烧和盗汗。1 个月后，您发现自己患有结核病。治疗两周后症状改善，两个月后症状消失。 | ○ | ○ | ○ |
| 隔离：出院后，您仍然具有传染性因此需要留在家中隔离。您至少需要 2 个月后才能回去工作。 | ○ | ○ | ○ |
| 治疗（6 个月）：持续 6 个月每天服用药物；首 2 个月服用 10 种药物，后 4 个月服用 4 种药物。您会出现恶心等副作用。您的家庭医生和护士会每天跟进您的用药情况。 | ○ | ○ | ○ |
| 对健康的持续影响：许多结核病患者康复后，仍感觉未能回复到之前的那么健康。 | ○ | ○ | ○ |
| 传染性：在您未被确诊和接受治疗之前，您可能会把病菌传染给与您接触的亲友。 | ○ | ○ | ○ |
| 羞耻: 其他人可能会发现您曾患有结核病。因为您缺席工作、被隔离，并且接触过您的同事和朋友也会被要求进行结核病感染检测，所以他们可能会猜测到您患有结核病。 | ○ | ○ | ○ |
| 死亡风险：有些人诊断得太晚。在加州，有 12% 的结核病患者死亡。 | ○ | ○ | ○ |

Intro_TB_top3 您最担心什么？请选择 3 项您担忧的影响。

*Display each choice if selected as “Very worrisome” or “Moderately worrisome”*

▢ 住院：您需要住院治疗两周。在医院里，您需要被隔离并与他人有极少接触。 (1)

▢ 健康状况不佳：经常咳嗽、体重减轻、发烧和盗汗。 1 个月后，您发现自己患有结核病。治疗两周后症状改善，两个月后症状消失。 (2)

▢ 隔离：出院后，您仍然具有传染性因此需要留在家中隔离。您至少需要 2 个月后才能回去工作。 (3)

▢ 治疗（6 个月）：持续 6 个月每天服用药物；首 2 个月服用 10 种药物，后 4 个月服用 4 种药物。您会出现恶心等副作用。您的家庭医生和护士会每天跟进您的用药情况。 (4)

▢ 对健康的持续影响：许多结核病患者康复后，仍感觉未能回复到之前的那么健康。 (5)

▢ 传染性：在您未被确诊和接受治疗之前，您可能会把病菌传染给与您接触的亲友。 (6)

▢ 羞耻: 其他人可能会发现您曾患有结核病。因为您缺席工作、被隔离，并且接触过您的同事和朋友也会被要求进行结核病感染检测，所以他们可能会猜测到您患有结核病。 (7)

▢ 死亡风险：有些人诊断得太晚。在加州，有 12% 的结核病患者死亡。 (8)

Intro_TB_ranking 您最担心什么？请拖放选项来进行排名（1：您最担忧，3：您最不担忧）。

*Display each choice if selected among top three*

_______ 住院：您需要住院治疗两周。在医院里，您需要被隔离并与他人有极少接触。 (1)

_______ 健康状况不佳：经常咳嗽、体重减轻、发烧和盗汗。 1 个月后，您发现自己患有结核病。治疗两周后症状改善，两个月后症状消失。 (2)

_______ 隔离：出院后，您仍然具有传染性因此需要留在家中隔离。您至少需要 2 个月后才能回去工作。 (3)

_______ 治疗（6 个月）：持续 6 个月每天服用药物；首 2 个月服用 10 种药物，后 4 个月服用 4 种药物。您会出现恶心等副作用。您的家庭医生和护士会每天跟进您的用药情况。 (4)

_______ 对健康的持续影响：许多结核病患者康复后，仍感觉未能回复到之前的那么健康。 (5)

_______ 传染性：在您未被确诊和接受治疗之前，您可能会把病菌传染给与您接触的亲友。 (6)

_______ 羞耻: 其他人可能会发现您曾患有结核病。因为您缺席工作、被隔离，并且接触过您的同事和朋友也会被要求进行结核病感染检测，所以他们可能会猜测到您患有结核病。 (7)

_______ 死亡风险：有些人诊断得太晚。在加州，有 12% 的结核病患者死亡。 (8)

Page Break

UCSF
University of California
San Francisco

Intro_LTBI_TPT
**潜伏性结核病**患者 (身体健康但感染结核病菌) 可能会也可能不会引致 **结核病**。一个人罹患**结核病** 的可能性取决于其年龄、吸烟状况、整体健康状况以及其它因素。美国疾病控制与预防中心 (CDC) 建议进行结核病预防性治疗。
但是，有些人会在预防结核病和 3 至 4 个月的治疗疗程间进行权衡 。更有可能患上**结核病**的人可能会从结核病预防性治疗中获益更多。对于不太可能患上**结核病**的人，即使接受结核病预防性治疗也收益不大，仅能进一步降低罹患结核病的可能性。我们想要了解您在患**结核病**的风险有多大的情况下才会采取预防性治疗。

F 对于下一个问题，请假设您患有**潜伏性结核病**。在未来 10 年内您有罹患**结核病**的风险。

*Randomize to BlockThreshold5 or BlockThreshold10, i.e., starting with 5% or 10% risk of TB disease. Only show one of the two blocks.*

**End of Block: BlockActiveTB**

---

**Start of Block: BlockThreshold5**

0.05 如果像您一样的 100 人中有 5 个人**在未来 10 年内会罹患结核病**，您是否愿意接受治疗？

- ○ 是，我愿意接受预防性治疗 (1)
- ○ 不，我不愿意接受治疗 (2)

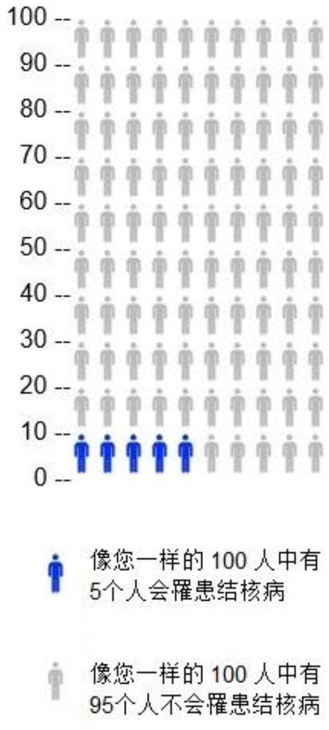


*If yes, skip to “less than 1 out of 100”. If no, continue to 50%. Follow the logic shown below. Arrows to the left mean “yes”, arrows to the right mean “no”. A straight arrow down to “stop” means go to the End of Block, regardless of the answer.*

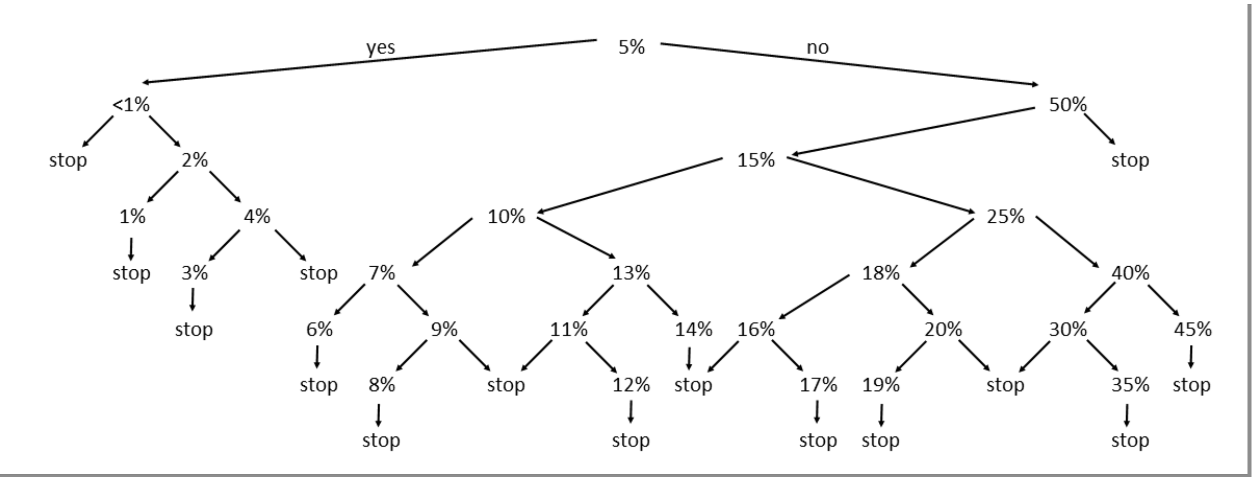


*At the end of this block, skip to Block_TBrisk_sliders.*

**End of Block: BlockThreshold5**

---

**Start of Block: BlockThreshold10**

T0.10 如果像您一样的 100 人中有 10 个人**在未来 10 年内会罹患结核病**，您是否愿意接受治疗？

- ○ 是，我愿意接受预防性治疗 (1)
- ○ 不，我不愿意接受治疗 (2)

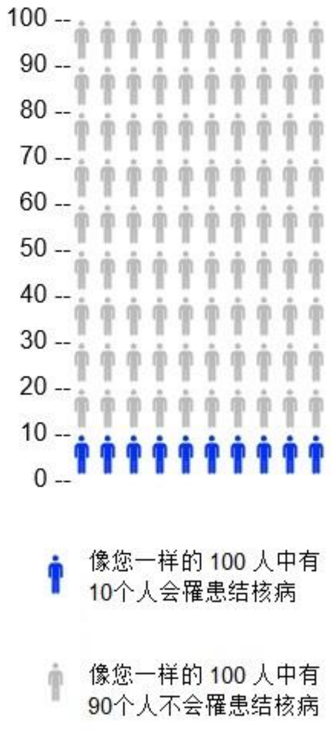

*If yes, skip to "less than 1 out of 100". If no, continue to 50 out of 100. Follow the logic shown below. Arrows to the left mean "yes", arrows to the right mean "no". A straight arrow down to "stop" means go to the End of Block, regardless of the answer.*

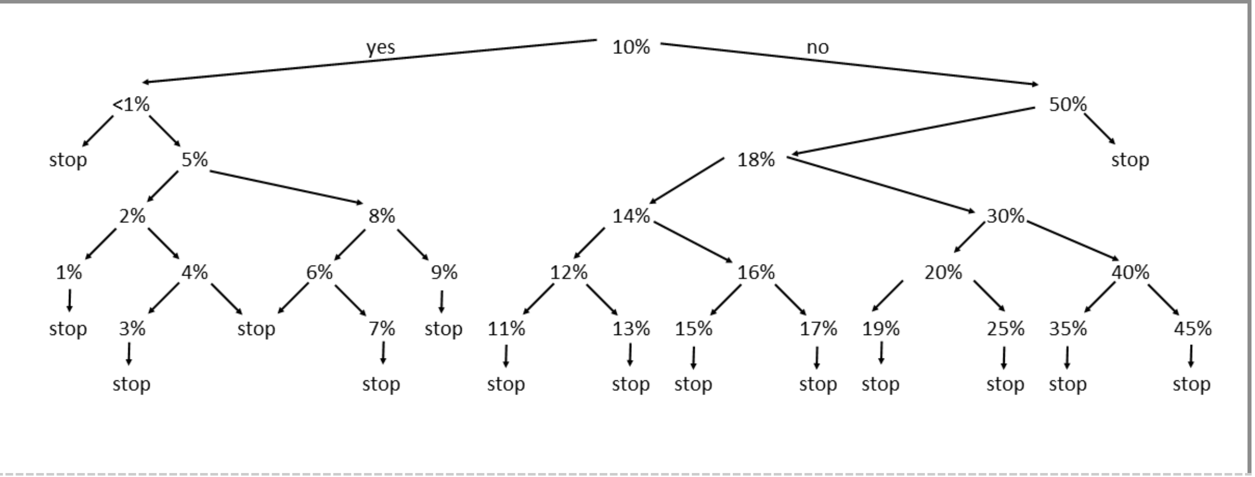


[We acknowledge that there was a translation error in the legend of the icon array for the risk below 1%. However, the question was translated and administered correctly, and we are confident the question was interpreted correctly by all participants.] Here is the figure legend with the error:]

T0.001 如果像您一样的 100 人中有少于 1 个人**在未来 10 年内会罹患**结核病，您是否愿意接受治疗？

- 是，我愿意接受预防性治疗 (1)
- 不，我不愿意接受治疗 (2)

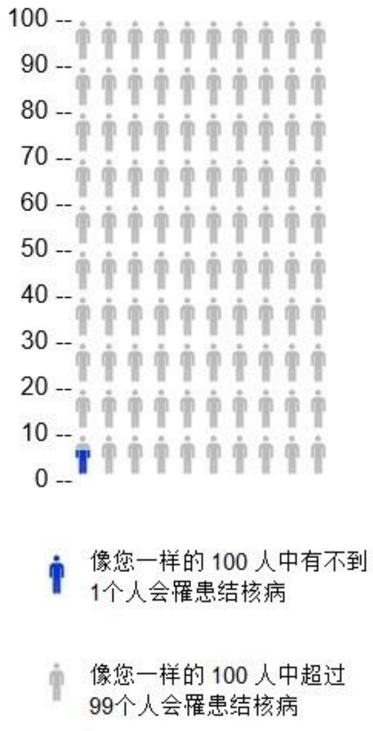


*At the end of this block, skip to Block_TBrisk_sliders.*

**End of Block: BlockThreshold10**

Start of Block: Block_TBrisk_sliders

TBsliders_intro 我们现在将讨论时间如何影响您愿意接受结核病预防治疗的想法。
[randomized order of questions: 10 years, 2 years, life time risk]

TBslider_10y
考虑一下您**在未来 10 年内罹患结核病**的概率。
要多高的患病概率，您才会考虑接受预防性治疗？（例如，想一下哪一种可能性较小？） 点击选择合适的答案。

| | 低概率（%） | 高概率（%） |
|---|---|---|
| | 0 10 20 30 40 50 60 70 80 90 100 | |
| **未来 10 年**罹患结核病的概率 (%) () | | |

TBslider_2y
假设您在**未来 2 年内**可能会罹患**结核病**。
要多高的患病概率，您才会考虑接受预防性治疗？

| | 低概率（%） | 高概率（%） |
|---|---|---|
| | 0 10 20 30 40 50 60 70 80 90 100 | |
| **未来 2 年**罹患结核病的概率 (%) () | | |

TBslider_lifetime 考虑一下您**终生罹患结核病**的概率。例如，假设您活到 90 岁，请考虑您在 90 岁之前罹患**结核病**的风险。 罹患**结核病**的概率要多高，您才会接受预防性治疗？

| | 低概率（%） | 高概率（%） |
|---|---|---|
| | 0 10 20 30 40 50 60 70 80 90 100 | |
| **终生**罹患结核病的概率 (%) () | | |

End of Block: Block_TBrisk_sliders

Start of Block: Block_TBrisk_sliders2

TBrisk_choice_time 您的医生向您解释罹患**结核病**的概率时，考虑到您自身罹患结核病的风险的情况下，什么时间范围对您来说最重要？

- ○ 1 年 (6)
- ○ 2 年 (1)
- ○ 5 年 (2)
- ○ 10 年 (3)
- ○ 15 年 (7)
- ○ 20 年 (8)
- ○ 50 年 (9)
- ○ 终生风险 (4)
- ○ 其它，请注明： (5) ________________________________________________

End of Block: Block_TBrisk_sliders2

---

Start of Block: Side effects

Intro_AEs
结核病预防性治疗有时可能会引起副作用，例如恶心或持续约一周的皮疹。这些副作用会自行消失。
多数患者无需看医生。

---

accept_AE 即使可能出现暂时的恶心或皮疹，您也会接受结核病预防性治疗吗？

- ○ 是 (1)
- ○ 可能 (2)
- ○ 否 (3)

---

accept_AE_0.3 每个人出现副作用（如恶心或持续约一周的皮疹）的概率是不同的。假设每 100 个像您一样的人中有 30 人出现恶心或皮疹。您还愿意接受结核病预防性治疗吗？

- ○ 是 (1)
- ○ 可能 (2)
- ○ 否 (3)

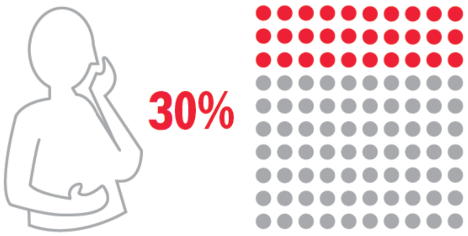


---

accept_AE_1.0 假设所有人都会感到恶心或出现持续约一周的皮疹。您还愿意接受结核病预防性治疗吗？

- 是 (1)
- 可能 (2)
- 否 (3)

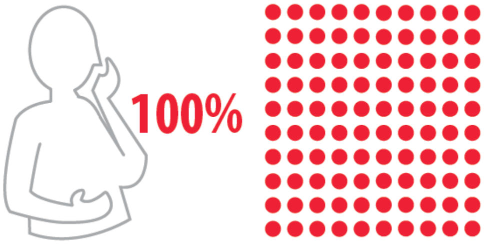


Page Break

Intro_Hepatotoxicity 结核病预防性治疗也可能会导致严重的副作用，例如肝功能损伤。每 100 人中有 0 到 5 人会出现肝功能损伤，出现这种副作用的人需要去医院就诊。

accept_AE_hepato 鉴于这种肝功能损伤概率（每 100 人中有 5 人），您还愿意接受结核病预防性治疗吗？

- 是 (1)
- 可能 (2)
- 否 (3)

value_AE
您更担心的是： 比较不常见的肝功能损伤（每 100 人中有 5 人会出现）或 比较常见的恶心或持续一周的皮疹（ 100 人中有 30 人会出现）？

- 我更担心肝功能损伤 (1)
- 我更担心恶心或皮疹 (2)
- 两者我都担心 (3)
- 两者我都不担心，预防结核病更重要。 (4)
- 其它，请注明： (5) ________________________________________________

accept_AE_open_2 针对结核病预防性治疗的副作用，您有什么意见？（没有则跳过此问题）

________________________________________________________________

End of Block: Side effects

Start of Block: Block_clinic_visit

reason_against_visit
接受结核病预防性治疗期间，我们建议您前往诊所就诊，以确保您感觉良好并帮助您完成治疗。
哪些因素会导致去诊所就诊变得困难？选择所有适用的选项。

- ▢ 就诊费用太昂贵（例如，没有保险或高额共付额）。 (10)
- ▢ 去诊所路途太长或交通过于复杂。 (8)
- ▢ 我想避免因前往诊所而产生的费用。 (1)
- ▢ 去诊所太费时间。 (2)
- ▢ 我去诊所时就会失去收入。 (4)
- ▢ 我去诊所就必须支付托儿费或类似费用。 (5)
- ▢ 我有其它要事（如工作或上学）。 (13)
- ▢ 我的身体状况受限，很难前往诊所。 (14)
- ▢ 我对去诊所就诊有顾虑（隐私问题、不信任医疗保健系统或负面的就诊经历）。 (15)
- ▢ 这对我来说不是优先考虑的事情，去诊所也不会有多大帮助。 (16)
- ▢ 其它，请注明： (9) ________________________________________________

---

reason_for_visits 在接受预防性治疗期间，促使您去诊所就诊的动力是什么？选择所有适用的选项。

- ▢ 我可以与家庭医生讨论可能出现的症状。 (6)
- ▢ 我的家庭医生可能有办法帮我避免副作用。 (12)
- ▢ 我可以与家庭医生讨论我的其它健康问题。 (5)
- ▢ 我的家庭医生能帮助我完成预防性治疗。 (4)
- ▢ 我对去诊所就诊感觉良好（例如：我信任医疗系统，或之前有良好的就诊经历）。 (13)
- ▢ 我可以获得帮助及健康知识，方便我管理自己的健康状况。 (14)
- ▢ 我的亲友建议我到诊所就诊。 (15)
- ▢ 去诊所就诊可以获得奖励或报销。 (16)
- ▢ 其它，请注明： (9) ________________________________________________

---

barrier_clinicvisit 如果您的家庭医生建议您在接受结核病预防性治疗期间到诊所就诊，您会接受并到诊所就诊吗？

- ○ 不，我不会接受或到诊所就诊。 (1)
- ○ 是，我愿意接受并到诊所进行**一次**就诊。 (2)
- ○ 是，我愿意接受并到诊所进行**多次**就诊。 (4)
- ○ 可能，我不确定。 (3)

---

ranking_visittype 您最希望通过何种方式与您的家庭医生进行每月一次的沟通？请拖放选项进行排名（1：首选，6：最后选择）

______ 与您的药剂师电话沟通 (1)
______ 与您的药剂师视频沟通 (6)
______ 亲自前往药房 (2)
______ 与您的家庭医生电话沟通 (3)
______ 与您的家庭医生视频沟通 (4)
______ 本人就诊 (5)

UCSF
University of California
San Francisco

End of Block: Block_clinic_visit

---

Start of Block: Block_blooddraw

Intro_blooddraw 医生可能会要求您每月抽血检查，以此监测您的肝肾是否健康，并评估您是否需要停止结核病预防性治疗。

---

barrier_bloodtests 您对于结核病预防性治疗期间的每月抽血检查有何看法？

- 如果必须抽血检查，我不会接受预防性治疗。 (1)
- 我仅会在开始预防性治疗之前做一次抽血检查。 (2)
- 我愿意接受每月一次的抽血检查，以确保我的肝肾处于健康状态。 (4)
- 我愿意接受结核病预防性治疗，但不愿接受抽血检查。 (5)
- 我不确定 (3)

End of Block: Block_blooddraw

---

Start of Block: Block_drugdruginteractions

Intro_ddi 大多数结核病预防性治疗会与其它药物及酒精会产生相互作用。

---

medications 您经常服用以下任意一种药物吗？勾选所有适用的项目。

- ▢ 您家庭医生开的药 (1)
- ▢ 非处方药 (2)
- ▢ 草药或其它传统药物 (3)

*Display this choice:If What is your sex? = Female*

- ▢ 避孕药或其他激素避孕药 (4)

*If Condition: Selected Count Is Equal to 0. Skip To: How often do you have a drink contain....*

---

stop_medications 假设您需要接受结核病预防性治疗，您愿意停用上述药物几个月吗？

- 是 (1)
- 否 (2)
- 可以停用一些药，但不能全停。 (4)
- 我不知道/不想回答 (3)

---

replace_medications 假设您需要接受结核病预防性治疗，您愿意尝试服用其它替代药物几个月吗？

- 是 (1)
- 否 (2)
- 可以替换一些药，但不能全换。 (4)
- 我不知道/不想回答 (3)

---

freq_alcohol 您喝酒的频率如何？

- ○ 从来不喝 (1)
- ○ 每个月喝一次或更少 (2)
- ○ 每月 2-4 次 (3)
- ○ 每周 2-3 次 (4)
- ○ 每周 4 次或更多 (5)

---

*Display this question: If How often do you drink alcohol? != Never*

stop_alcohol 假设您需要接受结核病预防性治疗，您愿意停酒几个月吗？

- ○ 是 (1)
- ○ 可能 (2)
- ○ 否 (3)
- ○ 我不知道/不想回答 (4)

**End of Block: Block_drugdruginteractions**

---

**Start of Block: Block_cost**

Intro_cost
结核病预防性治疗可能产生一些费用，例如：

共付额（您的保险不承保的部分）

交通费用（巴士票、出租车、停车费）

**照**顾亲属或孩子的费用（例如，去诊所时需要找其他人看护儿童）

---

threshold_cost 综合考虑上述所有费用，您愿意支付总额多少钱接受结核病预防性治疗？

- ○ $0，我只接受免费治疗 (1)
- ○ 最多$50 (2)
- ○ 最多$100 (3)
- ○ 最多$300 (4)
- ○ 大于$300 (5)

---

lost_income 除了额外支付治疗费用，您的收入也可能会减少（例如，您可能需要请假才能去就诊）。您愿意承担因接受**结核病**预防性治疗导致的收入损失是多少？

- ○ $0，我只会在不影响收入的情况下接受治疗 (1)
- ○ 最多$50 (2)
- ○ 最多$100 (3)
- ○ 最多$300 (4)
- ○ 大于$300 (5)

---

opportunity_cost 与寻求结核病预防性护理相比，您更愿意做什么？ （例如休闲活动或工作）

________________________________________________________________

---

University of California
San Francisco

opportunity_quant
您愿意为上述活动花多少钱（将时间花在您喜欢的活动上）？
（如果大于等于$300，请选择 $300。）

| | 0 | 30 | 60 | 90 | 120 | 150 | 180 | 210 | 240 | 270 | 300 |
| --- | --- | --- | --- | --- | --- | --- | --- | --- | --- | --- | --- |
| 以美元计算 () | | | | | | | | | | | |

End of Block: Block_cost

Start of Block: Block_reinfectionrisk

Intro_reinfection
结核病的预防性治疗只能治疗当前的结核病菌，但不能预防未来的感染。

reinf_travel 您前往**结核病**常见国家（例如亚洲、南美洲、中美洲、东欧、非洲）的频率是多少？

○ 每年都去 (1)
○ 每 2-5 年去一次 (2)
○ 每 6-10 年去一次 (3)
○ 每 10 年或更久才去一次 (4)
○ 我不打算前往这些国家 (5)

reinf_riskperceived
前往**结核病**常见国家旅行的人当中，大约每 100 人中有 1 人会感染（**潜伏性结核病**）。

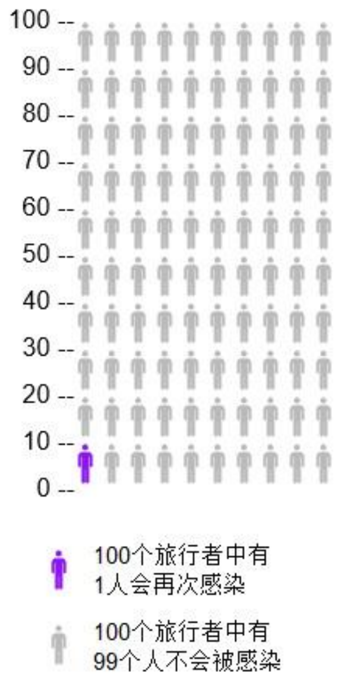


您会否担心完成预防性治疗后，会因旅行或其它原因再次感染结核病菌？

- ○ 一点也不担心 (1)
- ○ 有一点担心 (2)
- ○ 一般担心 (3)
- ○ 担心 (4)
- ○ 非常担心 (5)

---

reinf_open 您对于被感染的可能性是否有其它意见或看法？（没有则跳过）

________________________________________________________________

**End of Block: Block_reinfectionrisk**

---

**Start of Block: Intro_other**

Overall_rating 我们已经讨论了结核病预防性治疗的许多方面。当您在考虑是否接受结核病预防性治疗时，以下哪一项对您来说很重要？

| | 不重要 (1) | 一般重要 (2) | 非常重要 (3) |
|---|---|---|---|
| 如果不接受预防性治疗，您患结核病的可能性有多大。 (1) | ○ | ○ | ○ |
| 您出现恶心或皮疹的可能性有多大。 (2) | ○ | ○ | ○ |
| 您出现罕见但严重的肝损伤的可能性有多大。 (3) | ○ | ○ | ○ |
| 是否需要换药或停药几个月。 (4) | ○ | ○ | ○ |
| 是否需要停止饮酒几个月。 (5) | ○ | ○ | ○ |
| 去诊所就诊的频率。 (6) | ○ | ○ | ○ |
| 抽血检查的频率。 (7) | ○ | ○ | ○ |
| 可能产生的自付费用或收入损失。 (8) | ○ | ○ | ○ |
| 您再次感染结核病菌的可能性有多大。 (9) | ○ | ○ | ○ |

open_other 考虑结核病预防性治疗时，还有其它对您来说也很重要的因素吗？（如果没有则跳过。）

________________________________________________________________

End of Block: Intro_other

Start of Block: Block_demographics

civilstatus
完成今天的问卷调查之前，我们想要更加了解您。
您结婚了吗？

- ○ 已婚 (1)
- ○ 同居伴侣 (2)
- ○ 单身 (3)
- ○ 丧偶 (4)
- ○ 离异 (5)
- ○ 分居 (6)

---

householdsize 不包含您本人，您家里有多少人与您同住？

- ○ 家庭成员人数（不包含您本人）： (1) ________________________________________________

---

householdtype 哪些家庭成员住在您家里？勾选所有适用项。

- ▢ 子女（们） (1)
- ▢ 老人（们) (2)
- ▢ 患有影响免疫系统疾病（例如艾滋病毒）或正在服用免疫抑制剂（例如自身免疫性疾病）的人 (3)
- ▢ 以上全都没有 (4)

---

workstatus 您的就业状况是

- ○ 受雇 (1)
- ○ 自雇 (2)
- ○ 兼职 (3)
- ○ 未就业或正在找工作 (4)
- ○ 学生/延续教育 (5)
- ○ 退休 (6)
- ○ 其它，请注明： (7) ________________________________________________
- ○ 不愿回答 (8)

---

income 您的家庭年收入是多少（您家庭中每个人去年的收入总和）？

- ○ 低于$25'000 (1)
- ○ $25'001 至$50'000 (2)
- ○ $50'001 至$75'000 (3)
- ○ $75'001 至$100'000 (4)
- ○ $100'001 至$150'000 (5)
- ○ 高于$150'000 (6)
- ○ 不愿回答 (7)

---

healthinsurance 您有医疗保险吗？

- ○ 是 (1)
- ○ 否 (2)
- ○ 其它，请注明： (3) ________________________________________________

End of Block: Block_demographics

---

Start of Block: Block_surveyfeedback

diff_understand 您认为该问卷调查的问题很难理解吗？

- ○ 很难 (9)
- ○ 比较困难 (10)
- ○ 一般 (11)
- ○ 比较容易 (12)
- ○ 非常容易 (13)

---

diff_choose 您今天选择该问卷调查问题的答案时有多困难？

- ○ 很难 (11)
- ○ 比较困难 (12)
- ○ 一般 (13)
- ○ 比较容易 (14)
- ○ 非常容易 (15)

---

*Display this question:*
*If How difficult was it for you to understand the survey questions today? != Extremely easy*
*Or How difficult was it for you to choose answers to the survey questions today? != Extremely easy*

diff_open 您在理解或回答问题时遇到什么困难？

________________________________________________________________

---

survey_open
关于此次调查，您有何意见及建议吗？ （没有则跳过。）
点击“下一步”完成调查。

________________________________________________________________

End of Block: Block_surveyfeedback
